# Correlates of protection against symptomatic and asymptomatic SARS-CoV-2 infection

**DOI:** 10.1101/2021.06.21.21258528

**Authors:** Shuo Feng, Daniel J. Phillips, Thomas White, Homesh Sayal, Parvinder K. Aley, Sagida Bibi, Christina Dold, Michelle Fuskova, Sarah C. Gilbert, Ian Hirsch, Holly E. Humphries, Brett Jepson, Elizabeth J. Kelly, Emma Plested, Kathryn Shoemaker, Kelly M. Thomas, Johan Vekemans, Tonya L. Villafana, Teresa Lambe, Andrew J Pollard, Merryn Voysey, the Oxford COVID Vaccine Trial Group

## Abstract

**Background:** Although 6 COVID-19 vaccines have been approved by the World Health Organisation as of 16^th^ June 2021, global supply remains limited. An understanding of the immune response associated with protection could facilitate rapid licensure of new vaccines.

**Methods:** Data from a randomised efficacy trial of ChAdOx1 nCoV-19 (AZD1222) vaccine in the UK was analysed to determine the antibody levels associated with protection against SARS-CoV-2. Anti-spike and anti-RBD IgG by multiplex immunoassay, pseudovirus and live neutralising antibody at 28 days after the second dose were measured in infected and non-infected vaccine recipients. Weighted generalised additive models for binary data were applied to symptomatic and asymptomatic SARS-CoV-2 infection data from ChAdOx1 nCoV-19 recipients. Cubic spline smoothed log antibody levels, and weights were applied to account for potential selection bias in sample processing. Models were adjusted for baseline risk of exposure to SARS-CoV-2 infection.

**Results:** Higher levels of all immune markers were correlated with a reduced risk of symptomatic infection. Vaccine efficacy of 80% against primary symptomatic COVID-19 was achieved with an antibody level of 40923 (95% CI: 16748, 125017) and 63383 (95% CI: 16903, not computed (NC)) for anti-spike and anti-RBD, and 185 (95% CI: NC, NC) and 247 (95% CI: 101, NC) for pseudo- and live-neutralisation assays respectively. Antibody responses did not correlate with overall protection against asymptomatic infection.

**Conclusions:** Correlates of protection can be used to bridge to new populations using validated assays. The data can be used to extrapolate efficacy estimates for new vaccines where large efficacy trials cannot be conducted. More work is needed to assess correlates for emerging variants.

## Introduction

Within 17 months of the identification of SARS-CoV-2 in Wuhan, in response to the pandemic, a total of 6 COVID-19 vaccines have been recommended for use by the WHO as of 16^th^ June 2021.^1^ Vaccine efficacy of between 50% and 95% against symptomatic COVID-19 infections was reported using varying endpoint definitions.^2-7^ Real world evidence from vaccine rollout programmes has shown that COVID-19 vaccines are effective against severe disease, hospitalisation, and death, and reduce both asymptomatic infection and within household transmission.^8-13^

Global supply of COVID-19 vaccines remains limited despite massive production efforts. Authorization of new vaccines could help meet demand. As more countries implement vaccine programmes it will become increasingly difficult to conduct clinical efficacy studies of new vaccines. Understanding the relationship between immune responses to vaccines and protection against clinical outcomes is urgently needed to speed vaccine development. Knowledge of immune measures that are statistically associated with protection against disease (“correlates of protection”) may allow new vaccines to be authorised for use based on immunogenicity and safety data alone, when large efficacy trials are not feasible. In addition, understanding the immune response allows vaccines to be compared across cohorts of people who differ by age, race/ethnicity or other factors.

Both binding and neutralising antibodies are thought to be potential correlates of protection and are correlated with each other.^3,14-16^ Previous challenge studies of seasonal coronaviruses reported high levels of baseline neutralising antibody in uninfected or asymptomatic persons.^17^ However protection from infection with seasonal coronaviruses is not long lasting. ^17,18^

Early evidence from a cruise ship outbreak of SARS-CoV-2 suggested higher pre-existing neutralising antibodies were potential correlates of protection.^18,19^ A longitudinal cohort study of healthcare workers highlighted the association between baseline anti-spike and anti-nucleocapsid IgG and decreased risk of SARS-CoV-2 infection in the following 6 months.^19,20^

Evidence that antibodies may play a role in mediating protection against overt disease has come from vaccination and challenge studies in animals. Both neutralising antibody titres and Fc functional antibody responses correlate with protection induced by DNA and adenoviral vectored vaccines in this model. ^21,22^ Additionally, higher binding antibody levels from passively transferred monoclonal antibodies were more protective against re-challenge than lower levels of antibody.^21-24^

The ChAdOx1 nCoV-19 vaccine (AZD1222) is a chimpanzee adenoviral vectored vaccine with full length SARS-CoV-2 spike insert which was developed at the University of Oxford and is in widespread use globally produced by AstraZeneca and their manufacturing partners. We previously showed that estimates of vaccine efficacy against symptomatic COVID-19 infection were higher in subgroups with higher pseudovirus neutralisation antibody titres, or increased anti-spike IgG in vaccine clinical trials of ChAdOx1 nCoV-19 in adults.^3^ Here we report the relationship between the humoral immune responses to vaccination and protection afforded by this vaccine to facilitate further vaccine development.

## Methods

### Study description

The data included in this analysis comes from participants enrolled in COV002, a phase 2/3 randomised single blind vaccine efficacy trial conducted across 19 sites in the UK. A full description of the trial including immunogenicity, efficacy, and safety data, and the protocol has been previously published.^2,3,14,15,25^

Briefly, participants in the study were randomised to receive ChAdOx1 nCoV-19 or a MenACWY control vaccine. Efficacy cohorts (groups 4, 6, 9, 10) were randomised in a 1:1 ratio, immunogenicity cohorts were randomised in either a 5:1 (groups 2, 8, 5d), 3:1 (groups 1, 7), or a 1:1 ratio (group 5) depending on the group (see CONSORT diagram, Figure S1). A subset of participants received low dose vaccines for prime or boost doses in groups 1, 2, 4, and 5a (Figure S1). Open label groups and are not included in this report.

### Study endpoints and outcomes

Participants were reminded weekly to contact their study site if they experienced any of the primary symptoms of COVID-19 (fever ≥ 37.8°C; cough; shortness of breath; anosmia or ageusia) and were assessed in clinic, with a nose and throat swab taken for nucleic acid amplification testing (NAAT). Additionally, participants were asked to complete a nose and throat swab at home each week.

The outcomes for this analysis were 1) primary symptomatic COVID-19: a NAAT+ swab with at least one qualifying symptom, and 2) asymptomatic infections identified from weekly self-administered swabs and defined as a NAAT+ swab with no symptom reported. Sensitivity analysis of asymptomatic infections removed potential false-positive cases by restricting to those with higher viral load (Ct value < 30). NAAT+ participants who had symptoms other than the main five COVID-19 symptoms were categorised as non-primary symptomatic and were not included in correlates analysis.

Primary symptomatic COVID-19 outcomes were further classified according to whether a symptomatic participant reported shortness of breath or not, and whether 3 or more COVID-19 symptoms among 5 were present, indicators of more severe disease.

All endpoints were evaluated by a blinded independent clinical review committee.

### Immune markers and time points

A proportion of serum samples from vaccine recipients at the 28-day post-boost visit (PB28) were tested on three different assays with four assay readouts. All NAAT+ cases were tested if sample volume allowed, while a proportion of non-cases were tested. Samples were tested blinded to case status. The data from non-cases consisted mainly of the samples processed for the initial application for emergency use which needed 15% of samples included in the efficacy cohort to be processed on validated assays. We assume the mechanism of missingness for samples to be missing at random.^26^ To account for the missing data, factors associated with sample availability were controlled as weights in the analysis (see Correlates of risk below and Inverse probability weighting in Supplementary methods).

Anti-SARS-CoV-2 Spike and RBD IgG were measured by a multiplex immunoassay on the MSD platform at PPD. Antibody neutralisation was measured with a lentivirus-based pseudovirus particle expressing the SARS CoV-2 spike protein (Monogram) and by a normalised live microneutralisation assay (Public Health England).

Due to the limitations of laboratory capacity fewer samples were tested for virus neutralisation than were tested using the quicker multiplex assay.

### Study design and analysis populations

We first defined the Correlates Population by restricting to participants who met the eligibility criteria and received ChAdOx1 nCoV-19: participants were eligible for inclusion if they were baseline seronegative to the SARS-CoV-2 N protein at first vaccination, had their PB28 visit within a 14 to 42 day window after the second dose, and were followed up to at least 7 days after PB28 with no prior evidence of infection. Participants who received two doses were included in the analysis, either standard dose followed by standard dose (SDSD), or low dose followed by low or standard dose (LDSD or LDLD). 9 participants who received mixed schedules (one dose of ChAdOx1 nCoV-19 and one dose of MenACWY control) in error were excluded from analysis (Figure S1). The same eligibility criteria were applied to define a Control Population of MenACWY recipients.

Among the ChAdOx1 nCoV-19 Correlates Population, those who had biomarker data available comprised the Correlates Cohort. Participants who tested NAAT positive more than 7 days after PB28 were defined as cases while those who did not have a positive test were defined as non-cases. The 7 day window was implemented to exclude cases in which exposure is likely to have occurred before a blood sample was taken.

### Baseline exposure risk to SARS-CoV-2 infections

To control for potential confounding due to variation in exposure risk among participants with available immune marker data, a logistic regression risk model was developed among the Control Population of MenACWY recipients. Baseline factors associated with exposure risk were used to model the probability of being NAAT positive in this population. Baseline variables for the risk model included age in years, ethnicity (white and non-white), BMI (<30 kg/m2, ≥30 kg/m2), co-morbidities (having any of: respiratory disease; cardiovascular disease; or diabetes) and healthcare worker status (non-healthcare worker, healthcare worker exposed to no more than 1 COVID patient on an average day, healthcare worker exposed to 1 or more COVID patients on an average day). The linear predictor from the risk model developed using the MenACWY Control Population was used to predict the baseline risk of exposure in the ChAdOx1 nCoV-19 Correlates Cohort.

#### Correlates or risk (CoR)

The CoR analysis was conducted within the Correlates Cohort. Log-transformed immune marker values were analysed using generalised additive models (GAM) for binary data with a cubic spline smooth applied to immune marker values to allow a non-linear effect. The logit-transformed predicted baseline exposure risk was included as a linear covariate in the GAM model. A p value <0.05 from the approximate significance test from the smooth GAM was used to determine if an immune marker was associated with protection. Separate models were fitted for each immune marker controlling for baseline exposure risk, and weighted by inverse probability weights as described in the supplementary methods section.

#### Correlates of vaccine efficacy (CoVE)

For each outcome, to derive the relative risk (RR) and CoVE, an estimate of the absolute averaged predicted risk from the CoR model was computed. The averaged absolute risk was then compared to the overall risk among MenACWY Correlates Population, which was itself weighted by the randomisation ratio for study groups not randomised 1:1.

Vaccine efficacy (VE) was defined as 100% x (1 – RR). Mean estimate of VE at each level of antibody in the dataset, as well as 95% confidence intervals were calculated from 10,000 bootstrap samples.

Further analysis details are provided in the supplementary appendix along with the original trial statistical analysis plan (SAP) and the separate SAP developed for immune correlates analyses. The immune correlates SAP leant heavily on the methods proposed in the publicly available SAP by the Coronavirus Prevention Network (CoVPN) Biostatistics Team.^27^

## Results

Table 1 summarises baseline characteristics for the defined Correlates Population, Control Population, and Correlates Cohort by cases and non-cases status. Participants were followed-up for a median of 88 and 85 days counting from 7 days after the PB28 visit, among Correlates and Control Populations respectively. Among 4,369 Correlates Population participants, there were a total of 174 breakthrough NAAT+ cases. Data were available for at least one assay for 171/174 (98.3%) cases and 1404/4195 (33.5%) non-cases. Data were available for anti-spike and anti-RBD IgG from 1318 PB28 samples (163 cases and 1155 non-cases, Table S1). A smaller set of data was available for analysis for pseudovirus neutralisation titres (149 cases, 828 non-cases) and for live neutralisation (110 cases and 412 non-cases) (Table S1). Cases were younger, with 84.2% of cases being 18-55 years compared with 71.6% of non-cases, and more likely to be healthcare workers (62.0% of cases were healthcare workers compared with 57.5% of non-cases, Table 1).

**Table 1.**
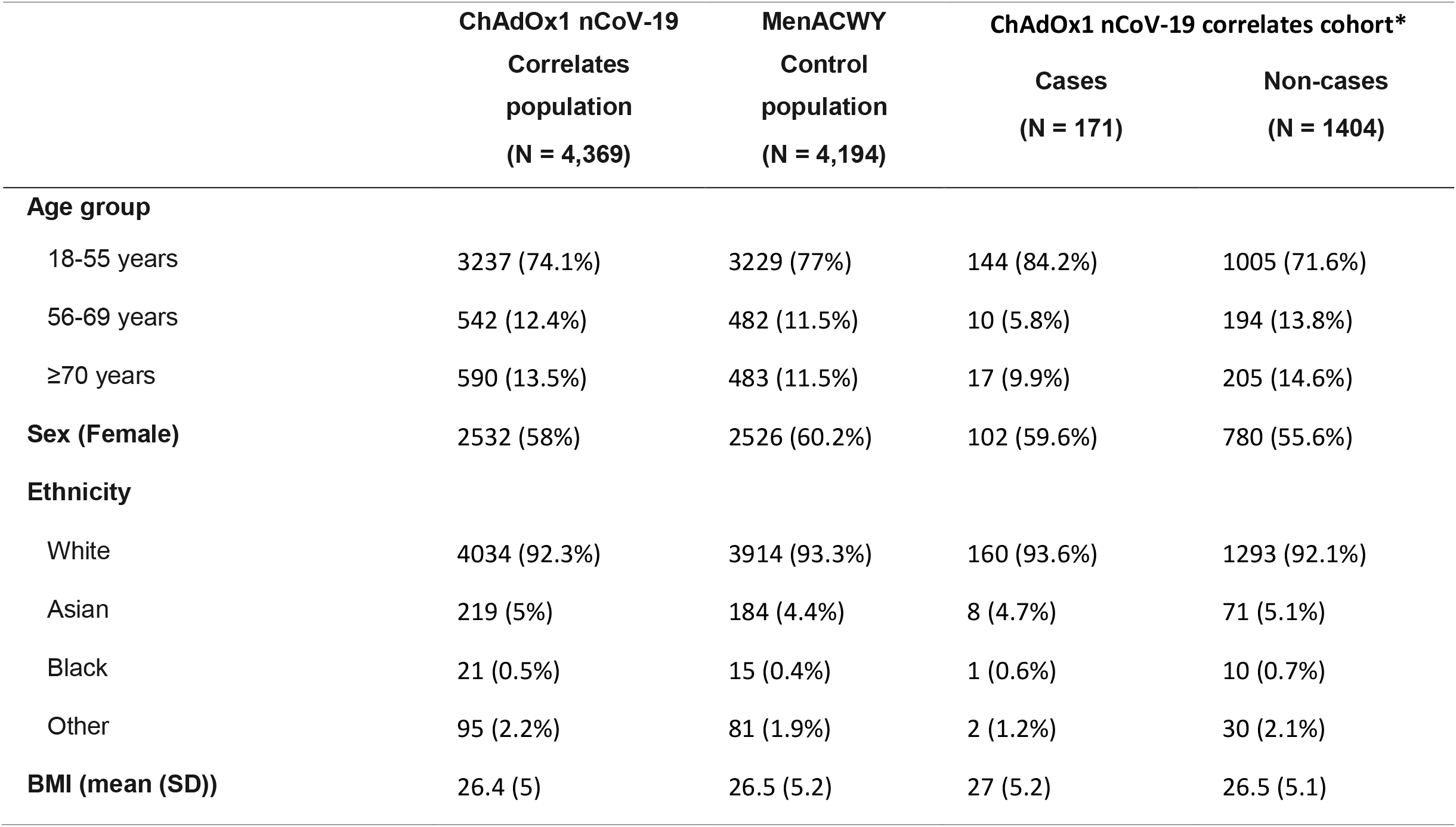
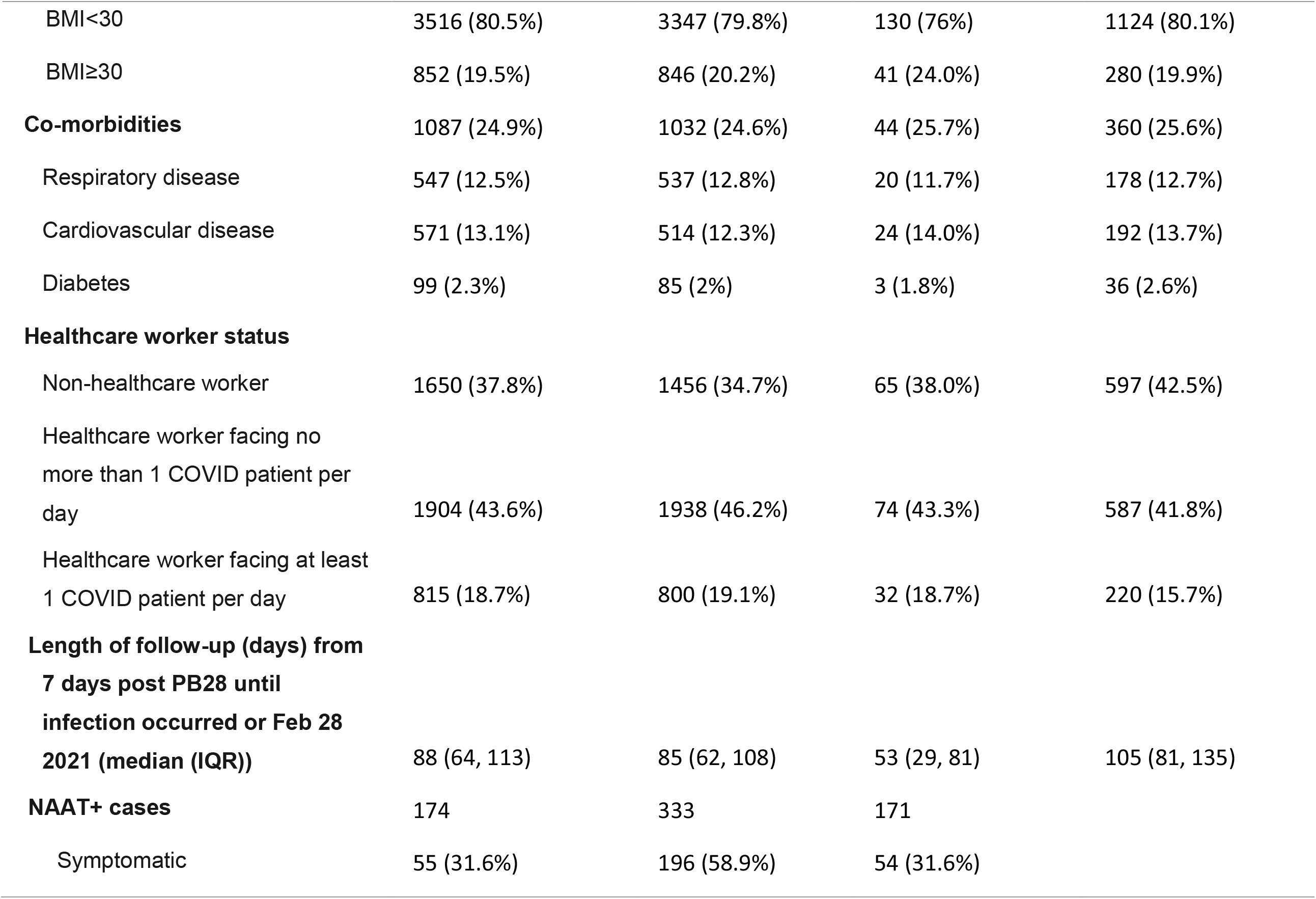
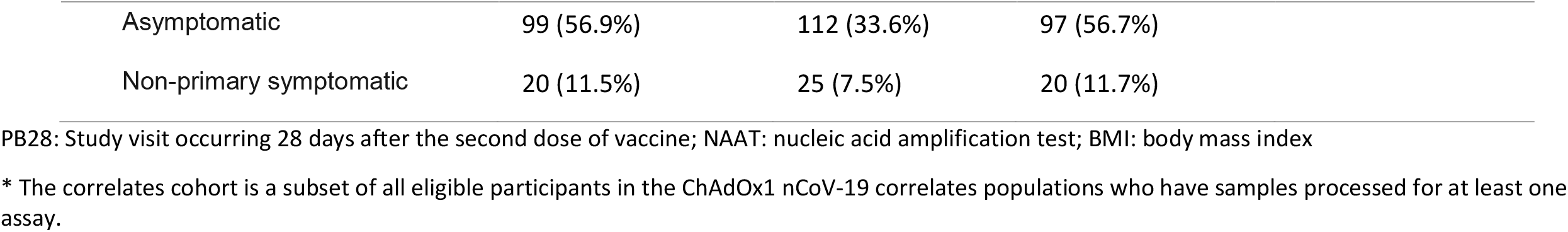
Baseline characteristics of correlates population, control population, and cases and non-cases among correlates cohort.

|  | ChAdOx1 nCoV-19<br>Correlates<br>population<br>(N = 4,369) | MenACWY<br>Control<br>population<br>(N = 4,194) | ChAdOx1 nCoV-19 correlates cohort* |  |
| --- | --- | --- | --- | --- |
|  |  |  | Cases<br>(N = 171) | Non-cases<br>(N = 1404) |
| **Age group** |  |  |  |  |
| 18-55 years | 3237 (74.1%) | 3229 (77%) | 144 (84.2%) | 1005 (71.6%) |
| 56-69 years | 542 (12.4%) | 482 (11.5%) | 10 (5.8%) | 194 (13.8%) |
| ≥70 years | 590 (13.5%) | 483 (11.5%) | 17 (9.9%) | 205 (14.6%) |
| **Sex (Female)** | 2532 (58%) | 2526 (60.2%) | 102 (59.6%) | 780 (55.6%) |
| **Ethnicity** |  |  |  |  |
| White | 4034 (92.3%) | 3914 (93.3%) | 160 (93.6%) | 1293 (92.1%) |
| Asian | 219 (5%) | 184 (4.4%) | 8 (4.7%) | 71 (5.1%) |
| Black | 21 (0.5%) | 15 (0.4%) | 1 (0.6%) | 10 (0.7%) |
| Other | 95 (2.2%) | 81 (1.9%) | 2 (1.2%) | 30 (2.1%) |
| **BMI (mean (SD))** | 26.4 (5) | 26.5 (5.2) | 27 (5.2) | 26.5 (5.1) |
| BMI<30 | 3516 (80.5%) | 3347 (79.8%) | 130 (76%) | 1124 (80.1%) |
| BMI≥30 | 852 (19.5%) | 846 (20.2%) | 41 (24.0%) | 280 (19.9%) |
| **Co-morbidities** | 1087 (24.9%) | 1032 (24.6%) | 44 (25.7%) | 360 (25.6%) |
| Respiratory disease | 547 (12.5%) | 537 (12.8%) | 20 (11.7%) | 178 (12.7%) |
| Cardiovascular disease | 571 (13.1%) | 514 (12.3%) | 24 (14.0%) | 192 (13.7%) |
| Diabetes | 99 (2.3%) | 85 (2%) | 3 (1.8%) | 36 (2.6%) |
| **Healthcare worker status** |  |  |  |  |
| Non-healthcare worker | 1650 (37.8%) | 1456 (34.7%) | 65 (38.0%) | 597 (42.5%) |
| Healthcare worker facing no more than 1 COVID patient per day | 1904 (43.6%) | 1938 (46.2%) | 74 (43.3%) | 587 (41.8%) |
| Healthcare worker facing at least 1 COVID patient per day | 815 (18.7%) | 800 (19.1%) | 32 (18.7%) | 220 (15.7%) |
| **Length of follow-up (days) from 7 days post PB28 until infection occurred or Feb 28 2021 (median (IQR))** | 88 (64, 113) | 85 (62, 108) | 53 (29, 81) | 105 (81, 135) |
| **NAAT+ cases** | 174 | 333 | 171 |  |
| Symptomatic | 55 (31.6%) | 196 (58.9%) | 54 (31.6%) |  |

|  |  |  |  |
| --- | --- | --- | --- |
| Asymptomatic | 99 (56.9%) | 112 (33.6%) | 97 (56.7%) |
| Non-primary symptomatic | 20 (11.5%) | 25 (7.5%) | 20 (11.7%) |

PB28: Study visit occurring 28 days after the second dose of vaccine; NAAT: nucleic acid amplification test; BMI: body mass index
\* The correlates cohort is a subset of all eligible participants in the ChAdOx1 nCoV-19 correlates populations who have samples processed for at least one assay.

Antibody levels at 28 days post-boost in cases and non-cases across four biomarkers are shown in Figure S2 (all p > 0.05). Anti-spike IgG and anti-RBD IgG were highly correlated with each other (Pearson correlation = 0.926) while the correlation between pseudovirus neutralisation titre and live neutralisation titre was modest (Pearson correlation = 0.572). Anti-spike IgG values were also correlated with pseudovirus neutralisation titres (Pearson correlation = 0.657) and live neutralisation titres (Pearson correlation = 0.600) (Figure S3).

The risk of symptomatic COVID-19 decreased with increasing levels of anti-spike IgG (p=0.003), anti-RBD IgG (p=0.018), pseudovirus neutralisation titre (p=0.005) and live neutralisation titre (p<0.001) (Figures 1A, Figure 1B, Table 2). In contrast, there were no significant associations between any of the assays and protection against asymptomatic infection including for sensitivity analysis restricting to high viral load (all p>0.05, Figure 2A, Table 2, Figure S4A, Figure S4B). When primary symptomatic COVID-19 cases were classified according to the presence of shortness of breath, we observed a similar trend with increasing immune marker levels associated with lower risk of infection (all p<0.05, Figure 3A, Figure S5A), but not for those with no shortness of breath (all p>0.05, Figure 3B, Table 2, Figure S5B). Higher pseudovirus and live neutralisation titre were associated with lower risk of infection for those who had 3 or more COVID symptoms (Figure S6A, Figure S6B).

**Figure 1A:**
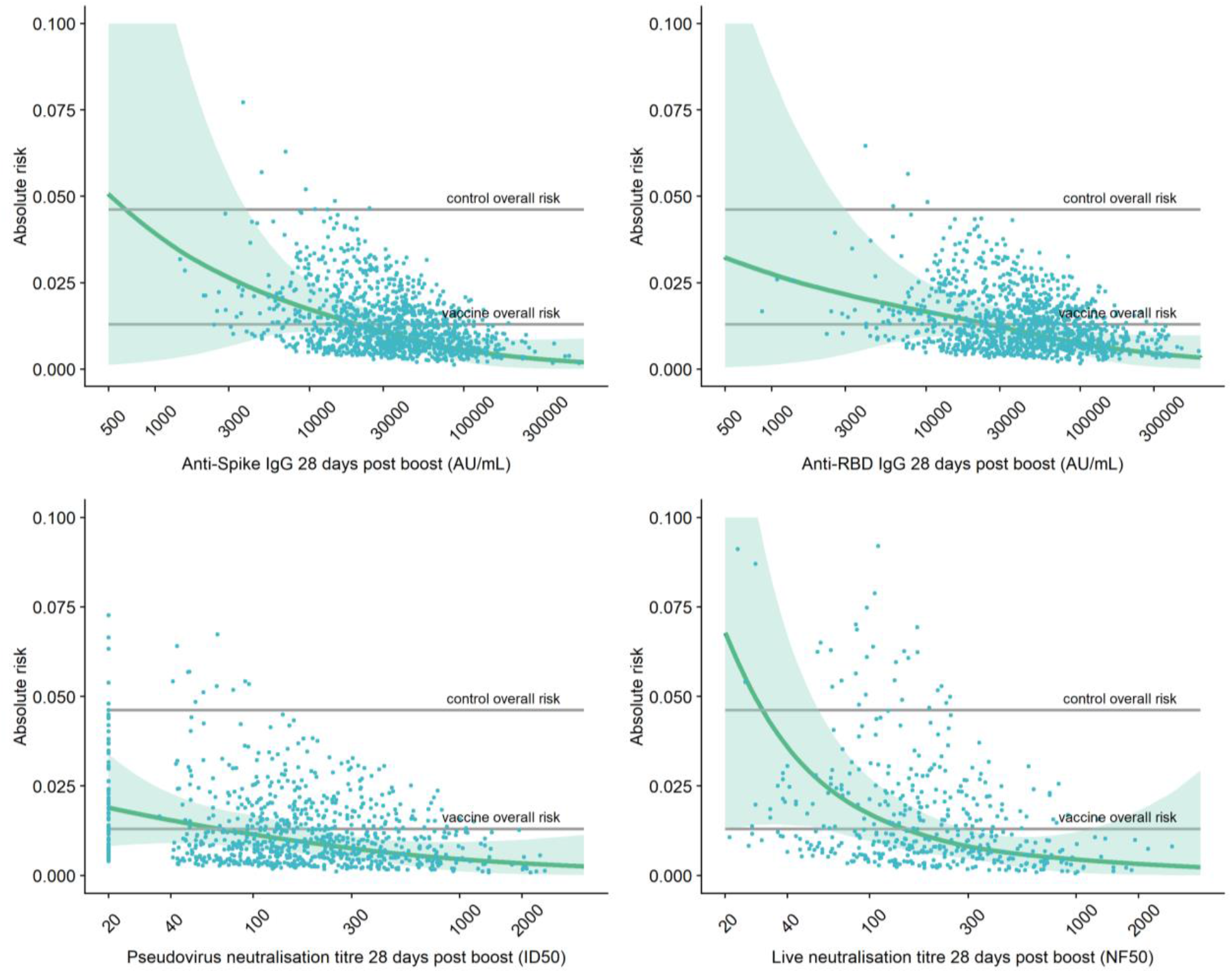
Adjusted risk of primary symptomatic COVID-19 as a function of immune markers measured 28 days post second dose. Top left: Anti-Spike IgG 28 days post boost Top right: Anti-RBD IgG 28 days post boost Bottom left: pseudovirus neutralisation antibody titres 28 days post boost Bottom right: live neutralisation antibody titres 28 days post boost. Grey lines show control (MenACWY) overall risk and vaccine (ChAdOx1 nCoV-19) overall risk. Blue dots show the absolute risk predicted from the model across the range of antibody values included in the analysis, adjusting for baseline exposure risk to SARS-CoV-2 infection (logit-transformed linear covariate including age, ethnicity, BMI, co-morbidities and healthcare worker status). Green shaded areas show the confidence interval around the predicted mean probability (green line)

**Figure 1B:**
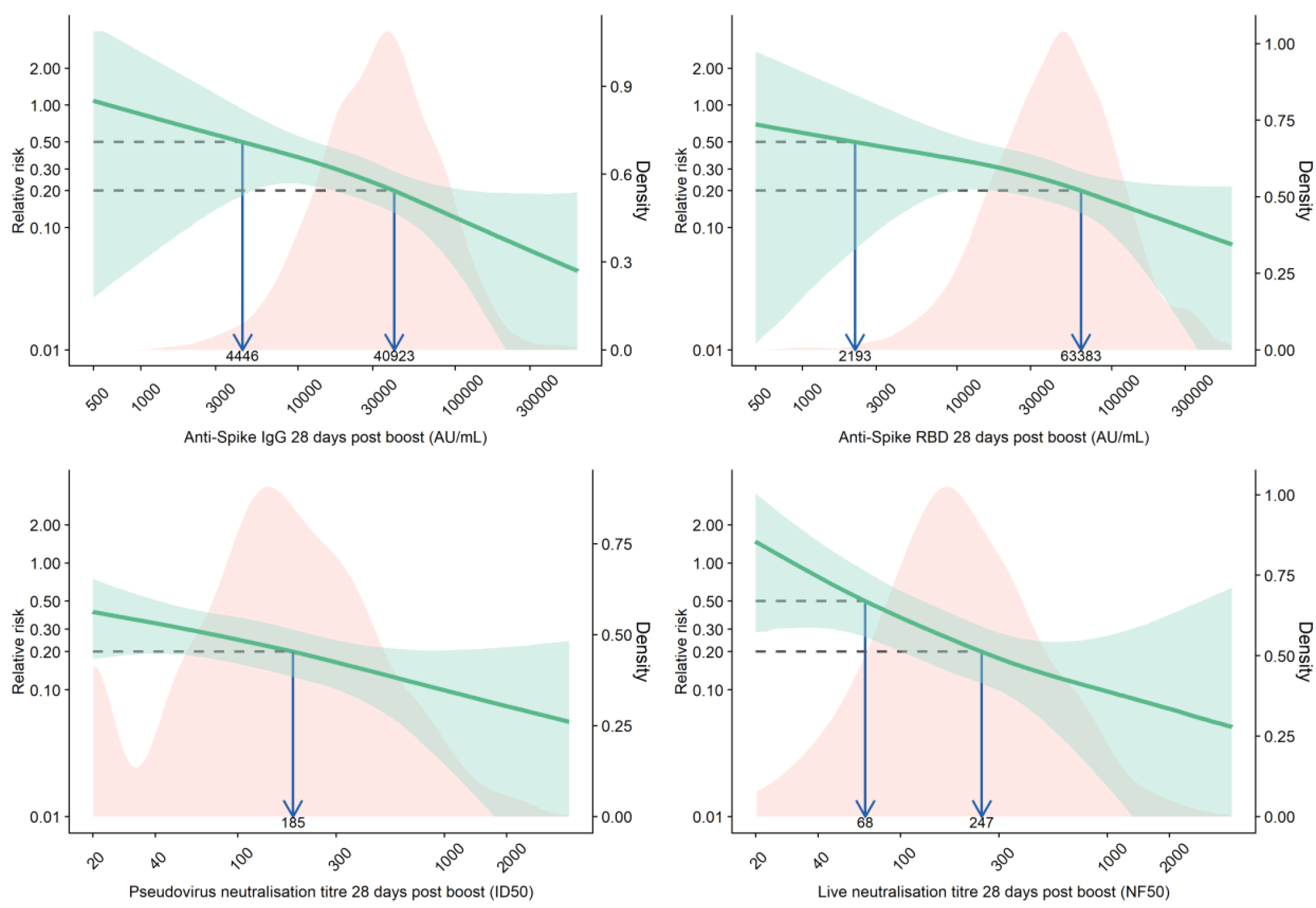
Relative risk of primary symptomatic COVID-19 among vaccine recipients compared with MenACWY control arm participants as a function of immune markers measured at day 28 post-second dose. Red shaded areas represent the immune marker density distribution. Green lines show the relative risk of infection among vaccine recipients compared to the MenACWY control arm participants. Green shaded areas are 95% bootstrapped confidence intervals for the relative risk. The arrows point to the immune marker values at 20% and 50% relative risk, i.e., 80% and 50% vaccine efficacy for illustrative purpose. The full range of VE estimates from 50 to 90% are shown in Table 2.

**Table 2.**
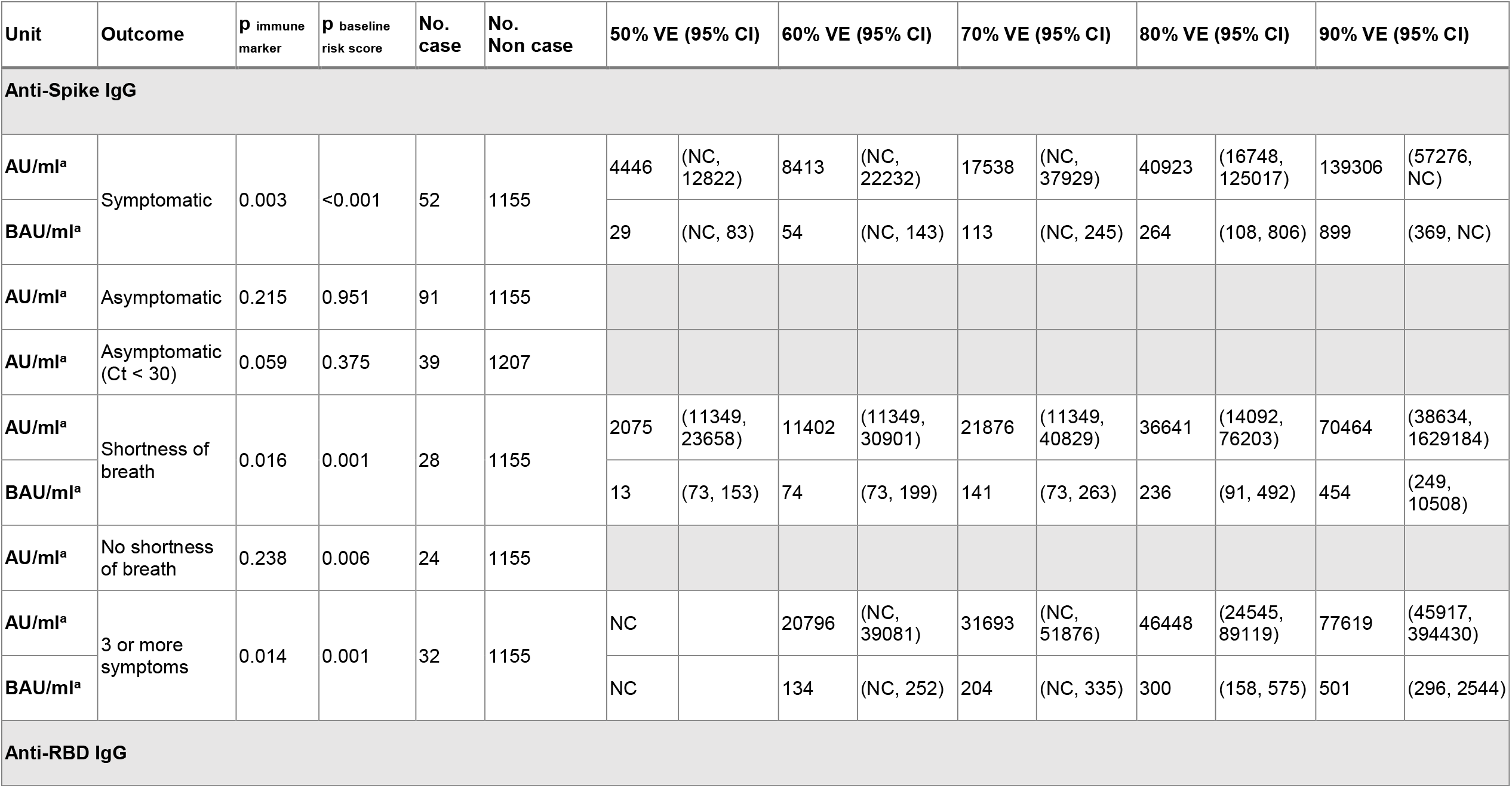
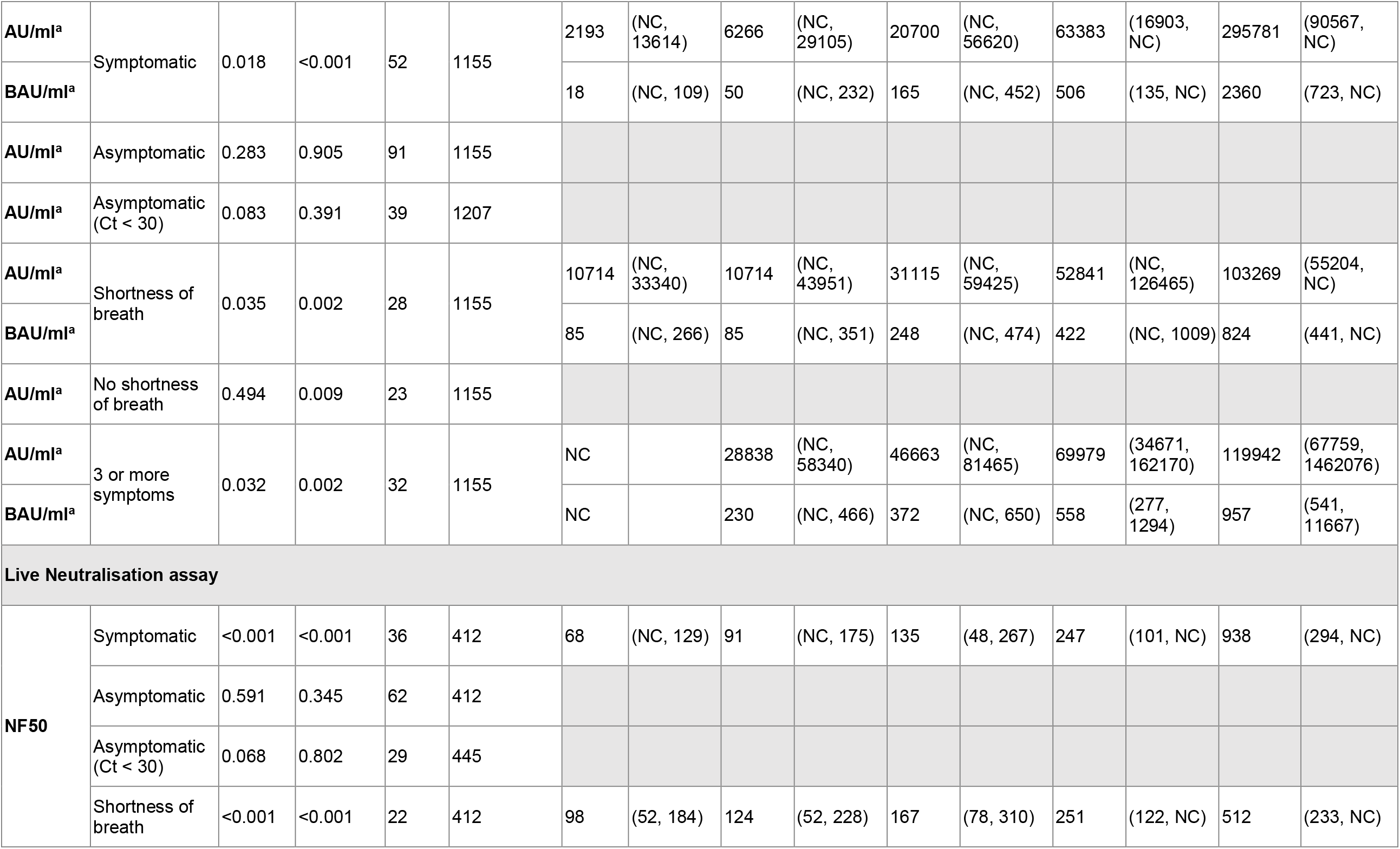
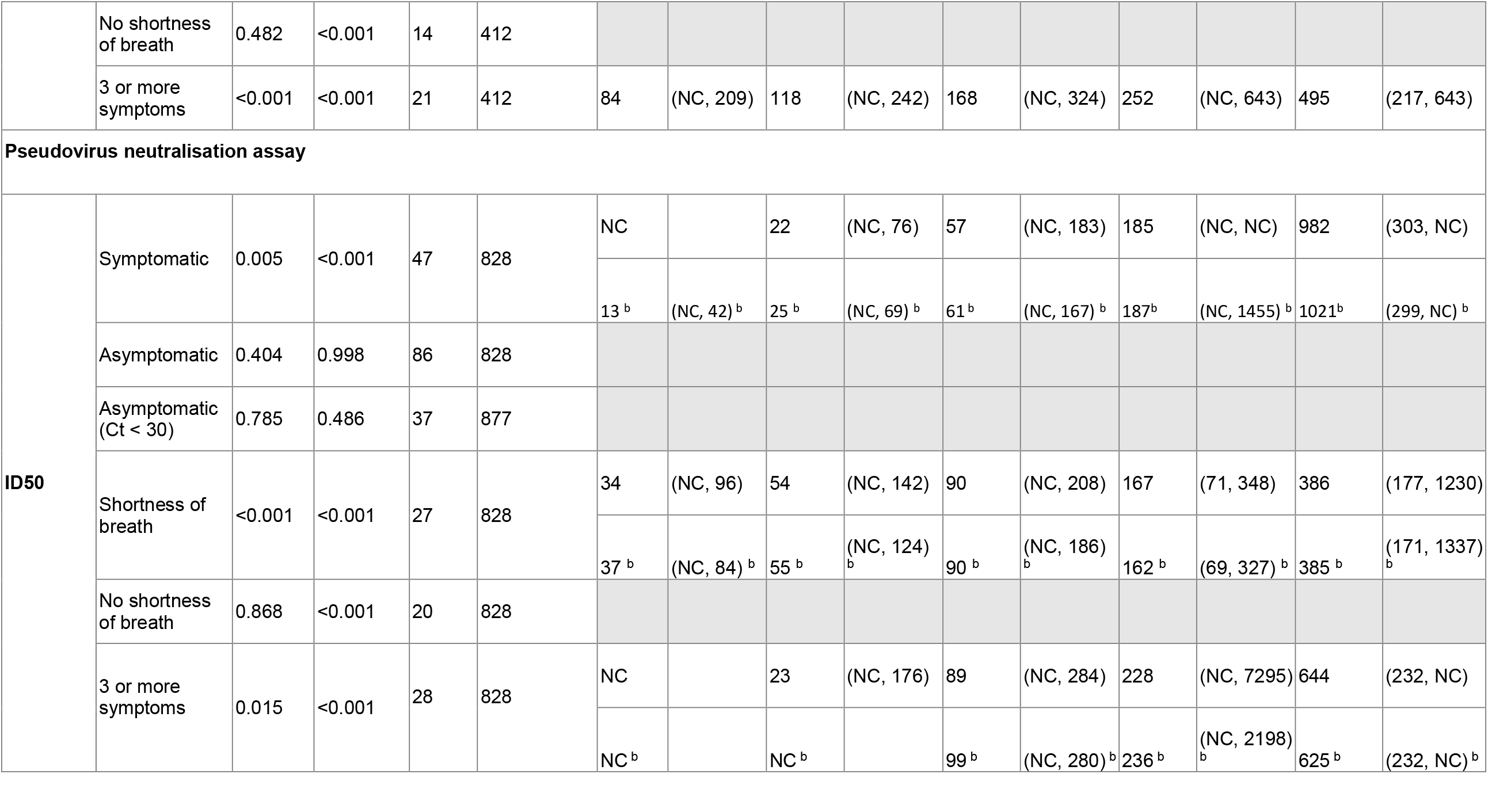
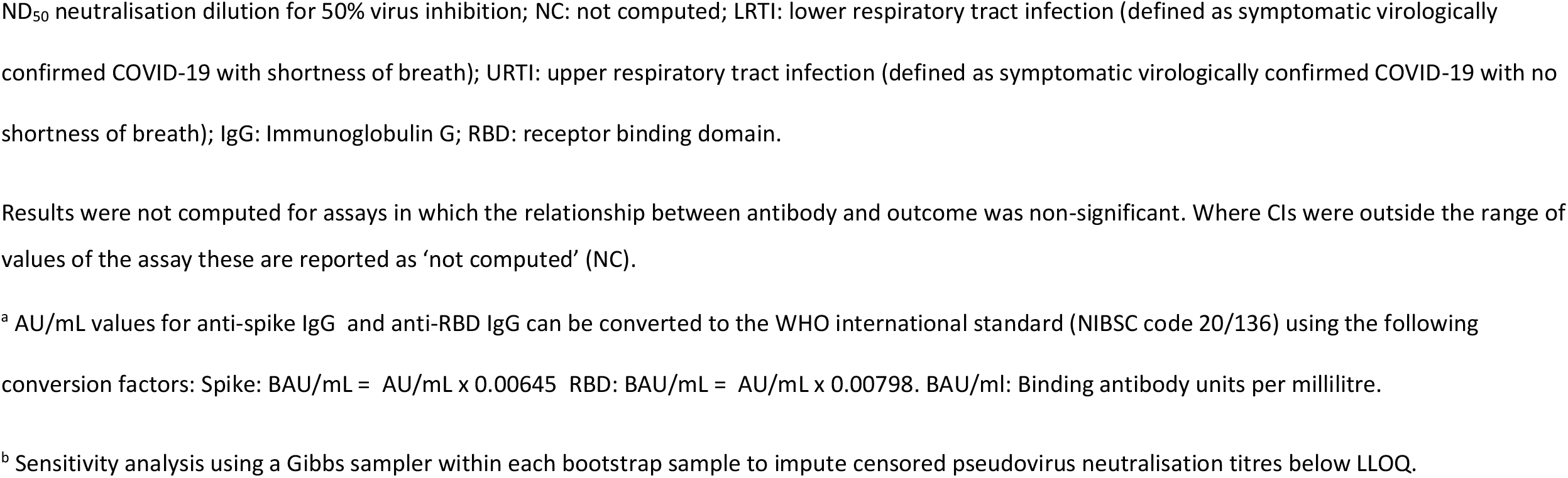
Outputs from generalised additive models, with immune marker values associated with 50%, 60%, 70%, 80% and 90% vaccine efficacy.

**Figure 1C:**
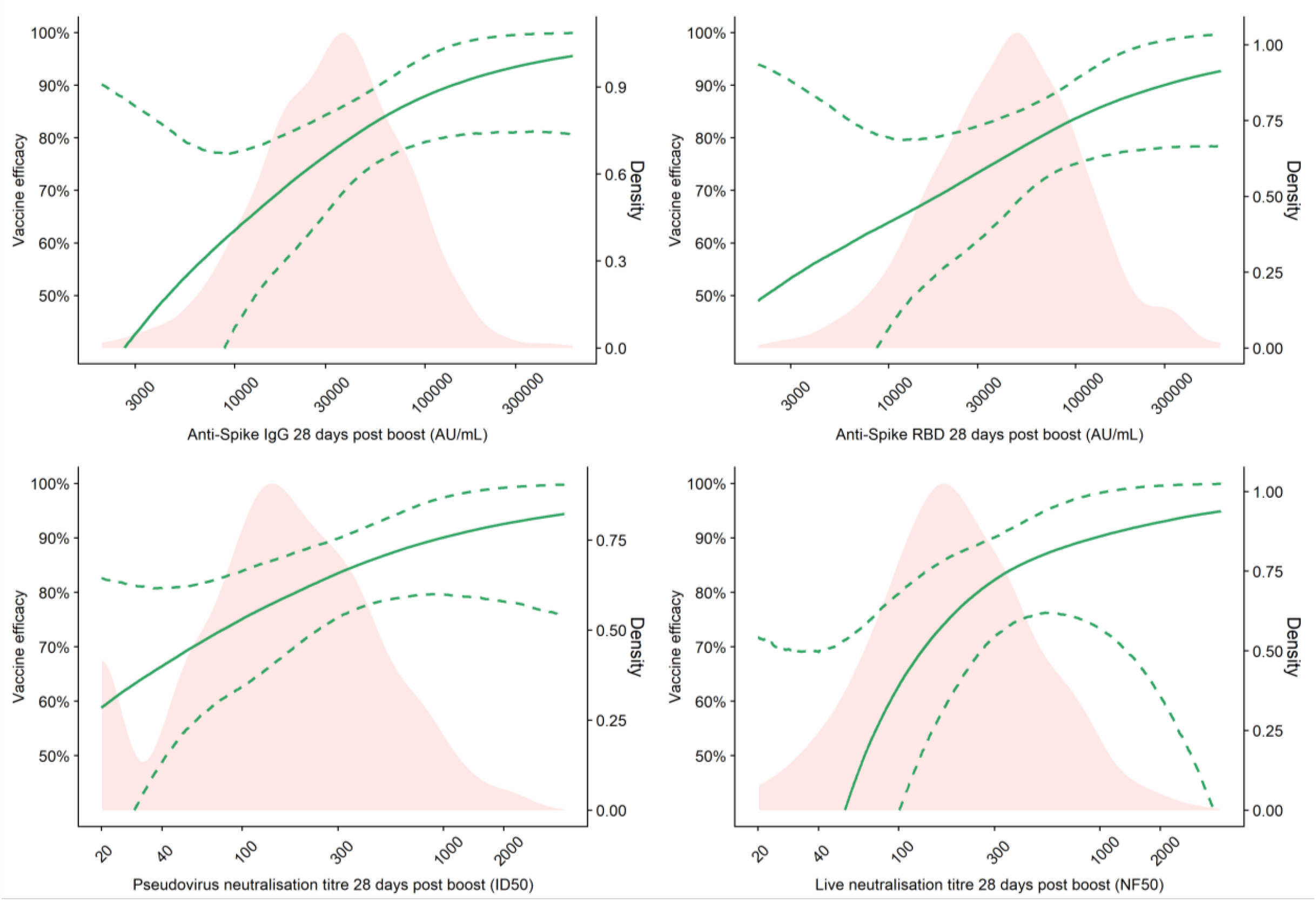
Vaccine efficacy against primary symptomatic COVID-19 as a function of immune markers measured at day 28 post-second dose. Red shaded areas represent the immune marker density distribution. Green lines show the vaccine efficacy and green dotted lines are 95% bootstrapped confidence intervals for vaccine efficacy.

**Figure 2A.**
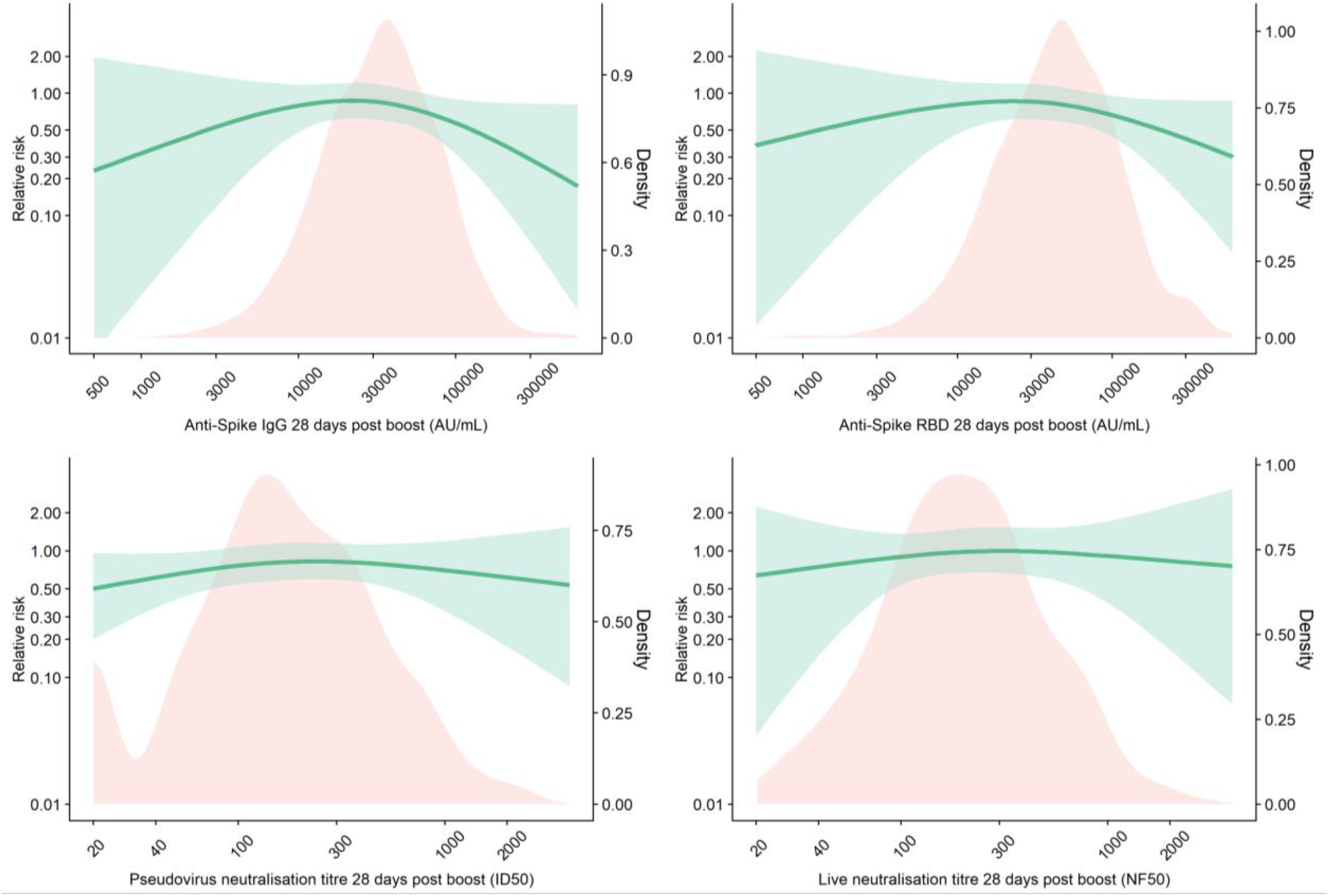
Relative risk of asymptomatic SARS-CoV-2 infection among vaccine recipients compared with the MenACWY control arm participants as a function of immune markers measured at 28 days post second dose. The red shaded areas represent the immune marker density distribution. Green lines show the relative risk of infection among vaccine recipients compared to the MenACWY control arm participants. Green shaded areas are bootstrapped 95% confidence intervals.

**Figure 2B.**
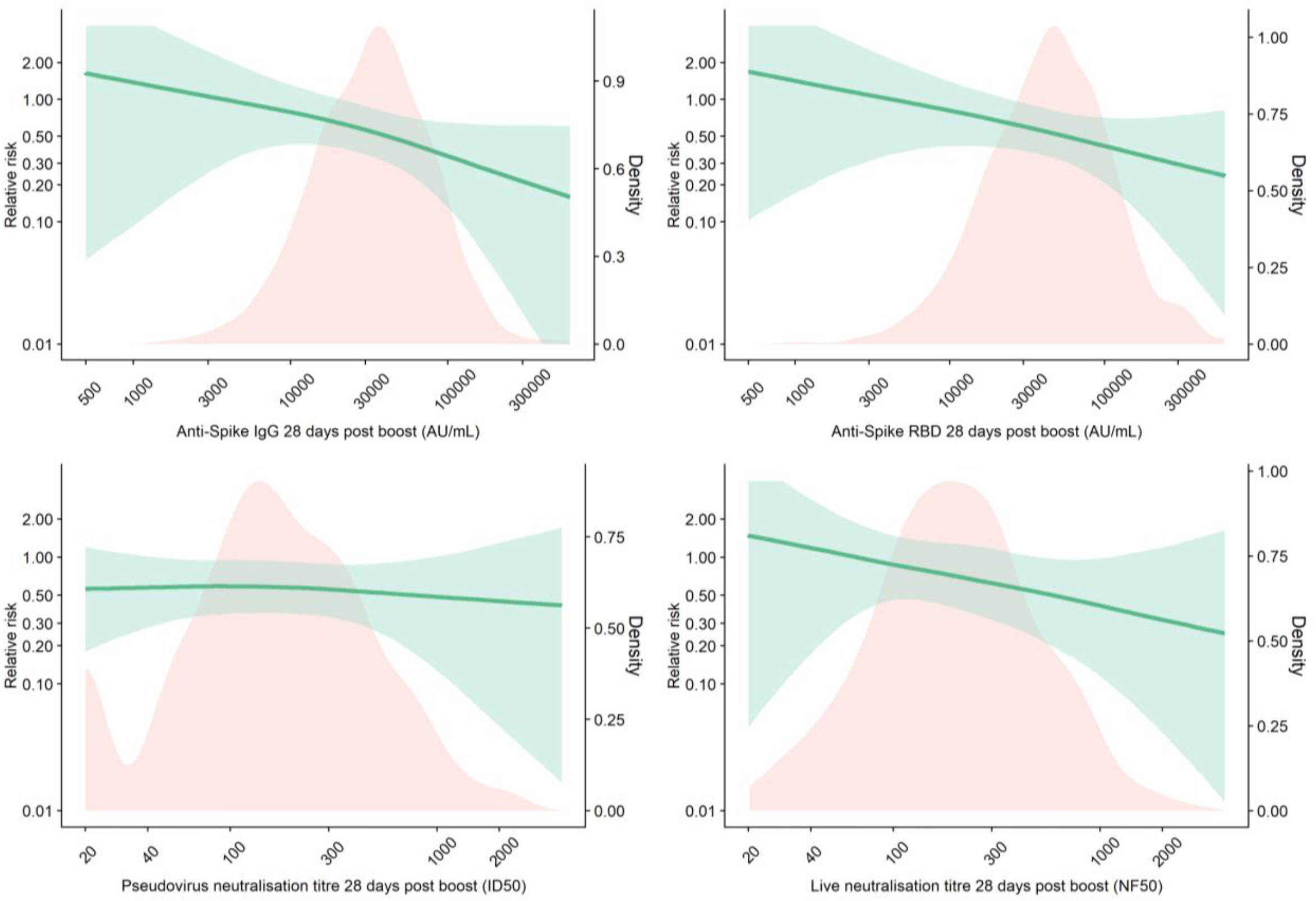
Sensitivity analysis showing relative risk of asymptomatic SARS-CoV-2 infection among vaccine arm compared with the control arm participants as a function of immune markers measured at 28 days post second dose excluding cases with low viral load (Ct ≥ 30) Red shaded areas represent the immune marker density distribution. Green lines show the relative risk of infection among vaccine recipients compared to the MenACWY control arm participants. Green shaded areas are 95% bootstrapped confidence intervals for the relative risk.

**Figure 3A.**
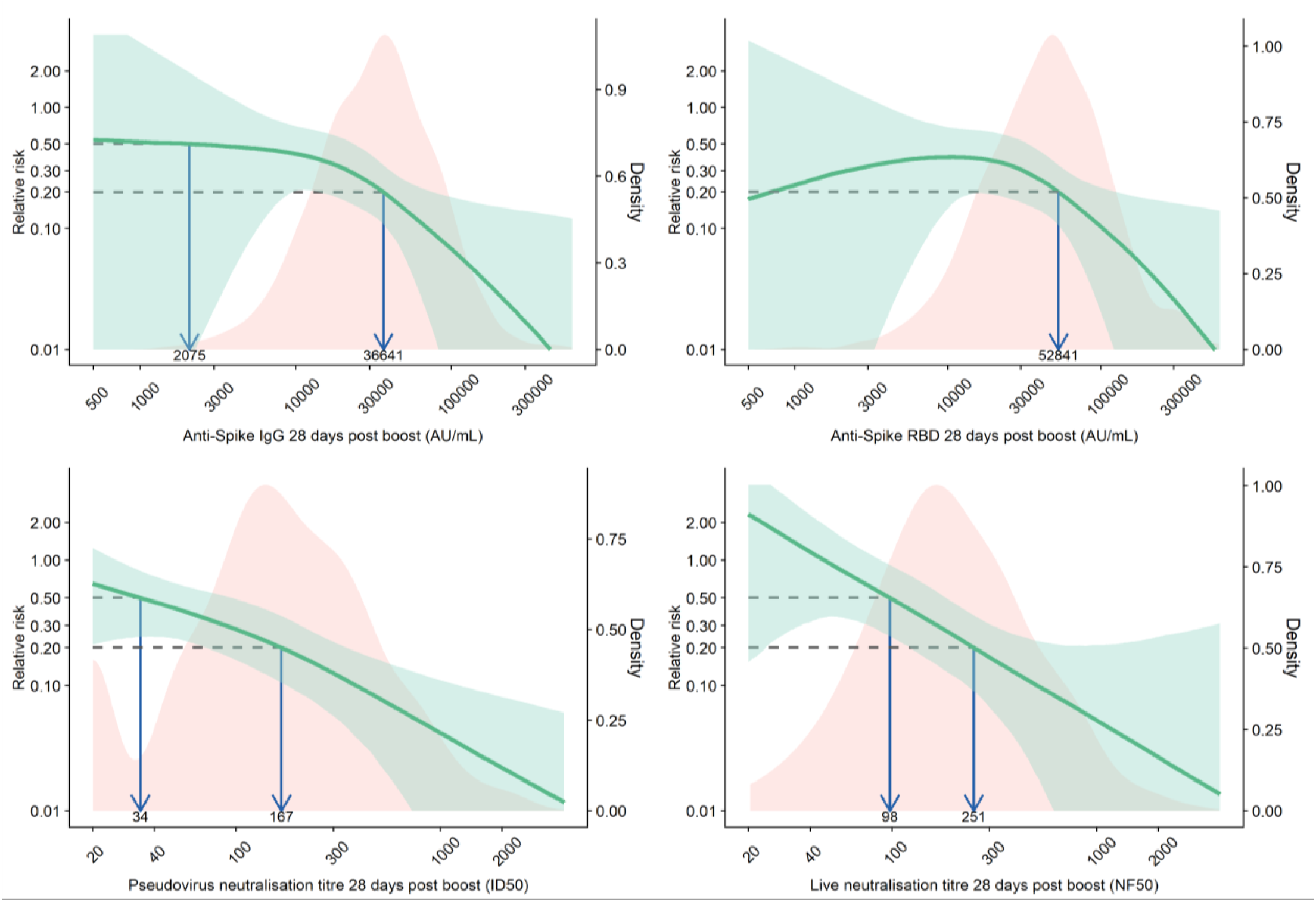
Relative risk of primary symptomatic SARS-CoV-2 infection with shortness of breath among vaccine recipients compared with MenACWY control arm participants as a function of immune markers measured at day 28 post-second dose. The red shaded areas represent the immune marker density distribution. Green lines show the relative risk of infection among vaccine arm compared to the control arm. Green shaded areas are 95% confidence intervals for the relative risk. The arrows point to the immune marker values at 20% and 50% relative risk, i.e., 80% and 50% vaccine efficacy.

**Figure 3B.**
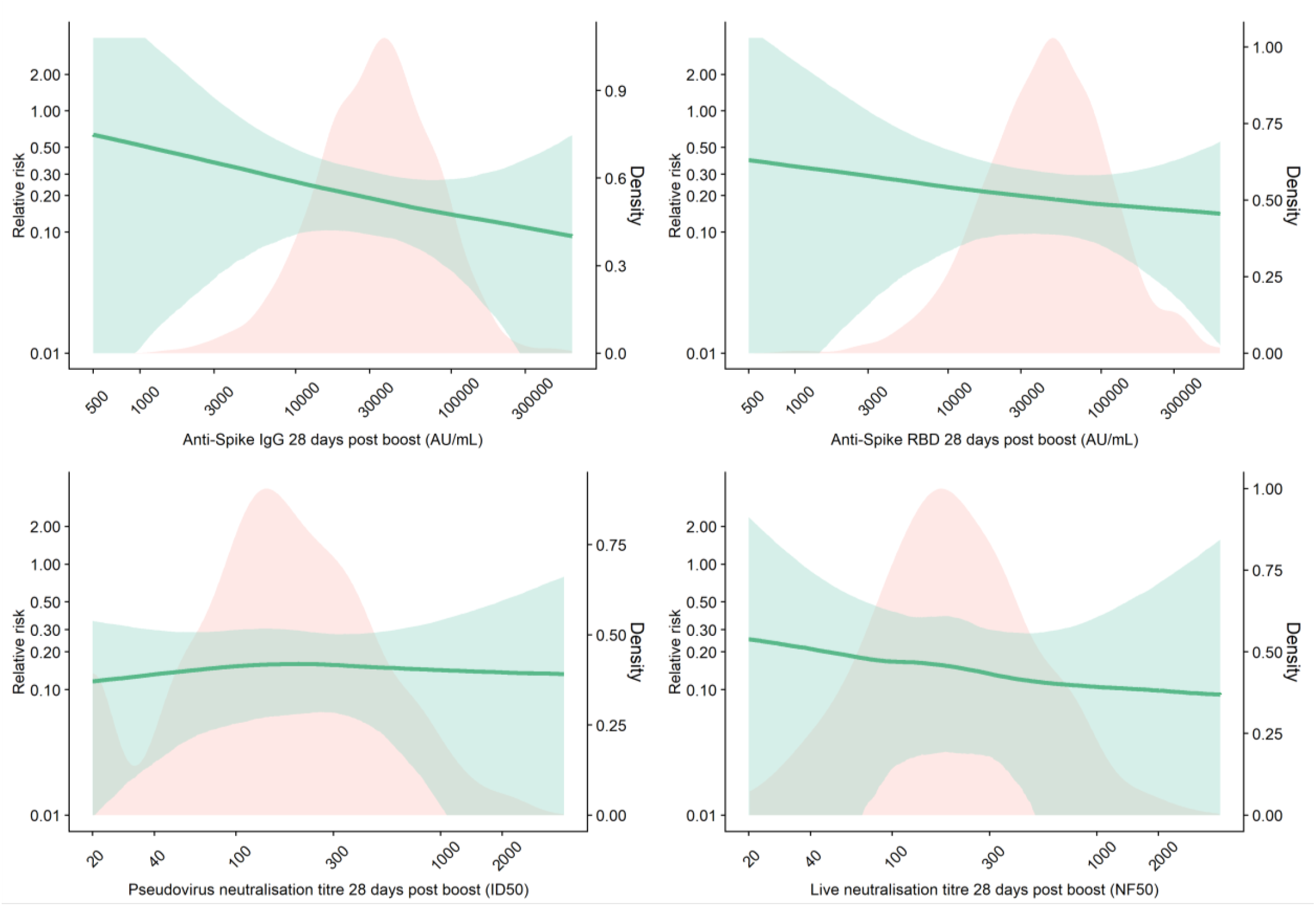
Relative risk of primary symptomatic SARS-CoV-2 infection with no reported shortness of breath among vaccine recipients compared with MenACWY control arm participants as a function of immune markers measured at day 28 post-second dose. The red shaded areas represent the immune marker density distribution. Green lines show the relative risk of infection among vaccine arm compared to the control arm. Green shaded areas are 95% confidence intervals for the relative risk.

The antibody level associated with 80% VE against primary symptomatic COVID-19, was 40923 (95% CI: 16748, 125017) arbitrary units (AU)/ml for anti-spike IgG, equivalent to 264 binding antibody units (BAU)/mL (95% CI 108, 806) using the WHO international standard (NIBSC code 20/136). For anti-RBD IgG 80% efficacy was achieved with median antibody of 63383 (95% CI: 16903, not computed (NC)) AU/mL (Figure 1B, Figure 1C, Table 2).

For pseudo and live neutralising antibody titres, neutralising titres at 185 (95% CI: NC, NC) and 247 (95% CI: 101, NC) respectively were associated with 80% VE against symptomatic infection. No values on these assays were associated with protection against asymptomatic infection (Table 2).

For all assays, when the analysis was restricted to symptomatic cases with shortness of breath, 80% VE was achieved at lower levels of immune markers than for symptomatic cases in general. Baseline exposure risk to SARS-CoV-2 infections was statistically significant in the GAM for all outcomes (all p < 0.05, Table 2) except for analyses of asymptomatic infections.

## Discussion

Here, we report an analysis of correlates of protection using data from 171 cases and 1404 non-cases, showing that higher anti-spike, anti-RBD IgG, and neutralising antibody titres are all associated with lower risk of symptomatic disease. We used immune responses in a phase 2/3 clinical trial to derive a model to predict absolute risk of infection, with appropriate adjustment for bias, assigning estimates for each level of antibody in the dataset. The relative risk of infection was then derived by reference to risk of infection in the control group. This is a robust approach to derive population estimates and adapted from recently described methods.^27,28^

The estimated anti-spike IgG level of 40923 AU/mL and the pseudo neutralising antibody titre of 185 associated with 80% VE in our models, were similar to the GMTs of 48961 AU/mL and 237.0 respectively, previously reported in a subgroup of participants vaccinated with ChAdOx1 nCoV-19 with a dose interval of at least 12 weeks between their 1^st^ and 2^nd^ dose – a regimen that provided 80·0% (95% CI 65·2 to 88·5) vaccine efficacy in the pooled analysis of data from the UK, Brazil and South Africa.^3^

In contrast, no serological measurements were shown to correlate with protection against asymptomatic infection or against symptomatic illness with only mild upper respiratory symptoms. This was unsurprising and is consistent with our interim combined VE analysis from UK and Brazil that vaccine efficacy against asymptomatic infection was 27.3% (−17.2 to 54.9) and was not significant at the 5% level.

Antibody correlates presented in this report, relate to protection against mild disease, defined as a PCR positive test with at least one symptom present. Weekly self-swabbing in the trial enabled detection of many mild cases. At these antibody titres, efficacy against more severe endpoints, used in other trials, would be higher than the estimates in this analysis. This has been confirmed in the analysis of real world effectiveness, in which the milder cases are not detected, after two doses of the vaccines in older adults in England where VE was 90% for Pfizer and 89% for ChAdOx1 nCoV-19 against symptomatic disease using the same case definition for both vaccines,^12^ while lower efficacy estimates were measured in our previously reported efficacy analysis with a milder disease endpoint.^2^

The correlates of vaccine efficacy reported here could be used to extrapolate efficacy to immunogenicity data for novel vaccines where clinical efficacy results are unlikely to be obtained. A trial of a new vaccine which produces antibody responses that are above the correlate values reported here, in at least 50% of participants (i.e. has a similar or higher median), might be expected to have similar efficacy against the clinical endpoints used in our UK trial, and higher efficacy against more severe endpoints. We provide correlates for vaccine efficacy estimates ranging from 50% to 90% to allow flexibility in the way these estimates are utilised by the regulators and policy-makers.

It has previously been shown that protection against lower respiratory tract infection (LRTI) may be easier to achieve than against upper respiratory tract infection (URTI) as challenge studies in rhesus macaques have shown stronger correlation between neutralising titres and the level of subgenomic mRNA in bronchoalveolar lavage samples than in nasal swab samples. ^29^ Similarly, ChAdOx1 nCoV-19 vaccinated hamsters, with low neutralising titres against B.1.351, were fully protected against LRTI following challenge with B.1.351, despite no evidence of protection against shedding of virus from the upper airway.^30^ Protection against upper respiratory tract or asymptomatic infections may be more closely associated with the presence of secretory IgA on the mucosal surface. ^31^

Interestingly, the vaccinated hamsters had complete protection against LRTI and reduced shedding in the upper airway after challenge with B.1.1.7, in the presence of neutralising antibody against this variant.^30^ These observations indicate that reduced neutralising capacity against B.1.351, and other variants of concern, might drive reduced protection against initial infection, and perhaps transmission, but protection against severe disease is maintained. Clinical trials of SARS-CoV-2 vaccines have consistently shown higher efficacy against more severe forms of disease such as hospitalisation or death, than against mild infections.^2-5,15,32^ We are unable to assess correlates of protection against severe disease or hospitalisation as there were no vaccinated participants hospitalised.

Although live- and pseudovirus neutralisation assays were modestly correlated with each other, the live virus assay was more closely associated with protection against symptomatic COVID-19 than the pseudovirus assay. This may reflect the sensitivity and dynamic range of the assays. Alternatively, this may reflect inherent differences between the assay systems, since pseudovirus neutralisation assays can express artificially high (or low) levels of target cell receptors which can in turn make it harder (or easier) to achieve neutralisation. Live neutralisation assays are accepted as the gold-standard methodology across fields and likely provide a more robust evaluation of neutralisation. Considering the high degree of variability between laboratories with neutralisation assays, binding antibodies would potentially provide more reliable, high-throughput and cost-effective results.

Protection against symptomatic COVID-19 is not absolute with any vaccine, and the results presented here show that there is no single threshold value for any of the assays investigated that is indicative of sterilising immunity. Instead, the probability of infection decreases on average with higher immune responses but substantial variation exists between individuals. This is similar to studies of respiratory syncytial virus where risk of infection decreased with higher antibody levels, although infections were still observed at high levels of antibody, suggesting a definitive individual threshold of protection does not exist.^33^ We provide antibody estimates that correspond with 50% to 90% VE however the wide confidence intervals around these estimates should be noted.

These estimates represent the antibody level observed 28 days after a *second* dose of vaccine that provide protection during the subsequent 4-6 month period among UK COV002 efficacy and immunogenicity cohorts. This is different from the antibody level that would protect an individual at the time of exposure to the virus. Further work is needed to determine the durability of antibody and long term protection after vaccination.

High levels of protection were noted after vaccination with one dose of a lipid nanoparticle RNA vaccine, despite modest levels of neutralising antibody, strongly supporting the concept that other mechanisms are at play as co-correlates of protection.^5,34^ We have previously shown that a wide range of Fc-mediated antibody functions are induced by vaccination, and it is possible that these functions may be important in the absence of neutralising antibody. ^35 35^ Furthermore, strong T cell responses induced by ChAdOx1 nCoV-19 may contribute to protection^14,16^ and have been associated with recovery from COVID-19 disease. ^36-38^ The relationship between antibody and T cell responses may differ depending on the type of vaccine used, and care should be taken in interpreting data from clinical testing of different vaccine technologies.

### Limitations

These analyses are based on cases of COVID-19 detected in a mainly white population in the UK, which were mostly due to B.1.177 and B.1.1.7 variants. In settings where these are not the dominant variants causing disease, or where neutralisation assays use different strains of the virus, the modelled relationships between immune markers and disease outcomes shown here may not apply. Furthermore, these analyses have been conducted on samples taken after 2 doses of ChAdOx1 nCoV-19 and might not apply to protection afforded by a single dose of the same vaccine or other COVID-19 vaccines. The potential role of T cells and interaction between humoral and cellular immunity has not been evaluated in this study.

### Conclusions

Correlates of protection can be used to bridge to new populations and new vaccines using validated assays. The data can be used to extrapolate efficacy estimates for new vaccines where efficacy data is unavailable.

## Data Availability

Anonymised participant data will be made available when the trial is complete, upon request directed to the corresponding author. Proposals will be reviewed and approved by the sponsor, investigator, and collaborators on the basis of scientific merit. After approval of a proposal, data can be shared through a secure online platform after signing a data access agreement. All data will be made available for a minimum of 5 years from the end of the trial.

## Acknowledgments

This Article reports independent research funded by UK Research and Innovation, Coalition for Epidemic Preparedness Innovations, and NIHR. We acknowledge support from Thames Valley and South Midland’s NIHR Clinical Research Network and the staff and resources of NIHR Southampton Clinical Research Facility and the NIHR Oxford Health Biomedical Research Centre. AJP is a NIHR senior investigator. We thank the volunteers who participated in this study.

We thank Professor Peter B. Gilbert and Dr Peter Dull for their advice and contributions to the methodology.

## Author Contributions

MV and SF designed the study. SF, DJP, TW, HS, BJ, KS, MV and IH contributed to the data analysis and methods. PKA, SB, CD, MF, BH, EP, TL and JV contributed to implementation of the study and/or laboratory experimentation. SF, MV, and AJP contributed to the preparation of the report. All authors critically reviewed and approved the final version.

