## Supplementary material for "Correlates of protection against symptomatic and asymptomatic SARS-CoV-2 infection": Figure S1, Figure S2, Figure S3, Figure S4A, Figure S4B, Figure S5A, Figure S5B, Figure S6A, Figure S6B, Table S1

### Supplementary Files

**Table of Contents**


1. Supplementary Methods……………………………………………………………………………………**Pg. 2**
2. Figure S1 Participant Flow chart showing inclusion in correlates models…………**Pg. 4**
3. Figure S2. Immune markers measured at day 28 post-second dose, in primary symptomatic, asymptomatic, non-primary cases, NAAT-positive cases and NAAT-negative non-cases……………………………………………………………………………………**Pg. 5**
4. Figure S3. Correlations between 1). top-left: Anti-SARS-CoV-2 Spike and RBD IgG, 2) top-right: Anti-SARS-CoV-2 Spike IgG and pseudovirus neutralisation titre, 3) bottom-left: Anti-SARS-CoV-2 Spike IgG and live neutralisation titre, 4) bottom-right pseudovirus neutralisation titre and live neutralisation titre…………………………**Pg. 6**
5. Figure S4A. Adjusted risk of asymptomatic SARS-CoV-2 infection as a function of immune markers measured 28 days post second dose……………………………………**Pg. 7**
6. Figure S4B. Sensitivity analysis showing adjusted risk of asymptomatic SARS-CoV-2 infection as a function of immune markers measured 28 days post second dose……………………………………………………………………………………………………**Pg. 8**
7. Figure S5A. Adjusted risk of primary symptomatic SARS-CoV-2 infection with shortness of breath as a function of immune markers measured 28 days post second dose……………………………………………………………………………………………………**Pg. 9**
8. Figure S5B. Adjusted risk of primary symptomatic SARS-CoV-2 infection with no shortness of breath as a function of immune markers measured 28 days post second dose……………………………………………………………………………………………………**Pg. 10**
9. Figure S6A. Adjusted risk of primary symptomatic SARS-CoV-2 infections with 3 or more COVID-19 symptoms as a function of immune markers measured 28 days post second dose………………………………………………………………………………………**Pg. 11**
10. Figure S6B. Relative risk of primary symptomatic SARS-CoV-2 infections with 3 or more COVID-19 symptoms among vaccine recipients compared with MenACWY control arm participants as a function of immune markers measured at day 28 post-second dose………………………………………………………………………………………**Pg. 12**
11. Table S1. Anti-SARS-CoV-2 Spike and RBD IgG, pseudovirus neutralisation antibody titres and live neutralisation antibody titres (Median and IQR) by primary symptomatic, asymptomatic/unknown, non-primary cases, NAAT positive cases and negative controls……………………………………………………………………………………**Pg. 13**
12. The Oxford Vaccine Trial Group……………………………………………………………**Pg. 14**
13. Acknowledgements……………………………………………………………………………**Pg. 16**
14. References……………………………………………………………………………………**Pg. 19**

**Supplementary methods**

#### Imputation on censored immune marker data in main analysis

Immune marker values were $\log_{10}$-transformed prior to analysis. Values which were censored at the lower limit of quantification (LLOQ) were imputed with the value LLOQ/2. Approximately 10% of the pseudovirus neutralisation titre were censored at the LLOQ, and sensitivity analysis were conducted by imputing these values using a Gibbs sampler.

#### Inverse probability weighting

Immune marker data were not available for everyone in the correlates population, and cases are over-represented in the immune marker datasets as these were preferentially processed over non-cases. Unadjusted estimates of absolute risk of infection will therefore be inflated and result in bias to correlates estimates. We used a logistic regression model to predict the probability that a participant will have immune marker data available to the analysis. The outcome variables were each immune marker, and predictors were age group (18-55 years, 56-69 years, 70 years or above), whether the participant is a case or non-case, the type of case (primary symptomatic, non-primary symptomatic, asymptomatic), prime-boost interval, and dosage (LD/LD, LD/SD, SD/SD). The inverse probability from this model was used to weight the correlates of risk models for each immune marker to remove this source of bias.

#### Bootstrap

We resampled from all participants enrolled in the study. For each bootstrap sample, we calculated the inverse probability weights to account for sampling bias. We then estimated the CoR by GAM, adjusting for the baseline risk exposure and weighting by inverse probability weights. We compared the predicted absolute risk from the GAM across the full range of antibody values, with the resampled MenACWY control population weighted overall risk. 10,000 bootstrap samples were used for each immune marker and outcome. The overall estimates for correlates of risk and correlates of vaccine efficacy were given by the median value in the bootstrap. 95% confidence intervals were calculated using the bootstrap percentile method, i.e., the 2.5% and 97.5% quantiles from the bootstrap.

Correlates and their CIs were not computed for assays in which the relationship between antibody and outcome was non-significant. Where CIs were outside the range of values of the assay these are reported as 'not computed' (NC).

#### Sensitivity analyses

##### Viral load

To account for potential of misclassification in asymptomatic infections, a sensitivity analysis was conducted excluding cases with lower viral loads (defined as those for whom all returned PCR positive tests had a Ct value ≥ 30) as these are potential false positives.

##### Imputation of censored antibody values

Approximately 10% of the pseudovirus neutralisation antibody titre were below the LLOQ. We performed a sensitivity analysis to account for the potential bias caused by imputing

LLOQ/2. Studies have shown that imputing LLOQ/2 can lead to bias and confidence intervals with poor coverage when a significant proportion of the data are censored[1-3]. When a bootstrap is required for missing data, Brand et al. (2019) found single imputation embedded inside a bootstrap showed better statistical properties than other methods[3]. We used an iterative Gibbs sampler proposed by Chen et al. (2013) to impute the censored log pseudoneutralising antibody values[2].

For each bootstrap sample we predicted the censored log pseudovirus neutralisation titre

based on the log anti-spike and anti-RBD IgG values, log live neutralising antibody values, baseline risk score and all variables used in the inverse probability weighting model.

More specifically, for each bootstrap sample we used a Bayesian imputation method to predict the censored values. Not all participants with results from the pseudovirus neutralisation titre also have results from the anti-spike, anti-RBD and live neutralising antibody titres. We iteratively predicted the missing and censored values for each antibody titre in a Gibbs sampler, constraining the predictions for the censored values to be less than or equal to the LLOQ. Each antibody titre was predicted by a Bayesian linear regression, with independent variables being the current prediction for all other titres and the other predictor variables in the model. We imputed a single value for each of the censored log pseudovirus neutralisation antibody values from the 100$^{th}$ iteration of the Gibbs sampler. The sensitivity analysis was then run on the imputed dataset for the bootstrap sample.

We initialised the Gibbs sampler by predicting the missing and censored values from a series of linear regressions on the non-missing data. We ran multiple chains on bootstrap samples and tested for convergence by inspecting trace plots of the censored log pseudovirus neutralisation titres. From these plots we determined the 100$^{th}$ iteration to be approximately converged.

#### Data cut-off

The data cut-off date for inclusion in this analysis was Feb 28, 2021.

#### Software

Data analysis was done using R version 3.6.1 or later.[4] The GAM was coded using the mgcv package.[5] Three knots were used for each GAM, and the smoothing parameter was estimated by generalized cross validation.

#### Figure S1 Participant Flow chart showing inclusion in correlates models

**Single blind immunogenicity and efficacy cohorts receiving ChAdOx1 n-CoV 19**

| **Group 1** | **Group 2** | **Group 4** | **Group 5** | **Group 6** | **Group 7** | **Group 8** | **Group 9** | **Group 10** |
|---|---|---|---|---|---|---|---|---|
| Age 56-69 years<br>LD N=0<br>LDLD N=30<br>LDSD N=30 | Age 70+ years<br>LD N=8<br>LDLD N=46<br>LDSD N=46 | Age 18-55 years<br>LD N=242<br>LDLD N=51<br>LDSD N=1,424 | Age 18-55 years<br>LD N=2<br>SD N=46<br>LDSD N=63<br>SDSD N=64 | Age 18-55 years<br>SD N=325<br>SDSD N=1,973 | Age 56-69 years<br>SD N=30<br>SDSD N=30 | Age 70+ years<br>SD N=51<br>SDSD N=49 | Age 56-69 years<br>SD N=8<br>SDSD N=505 | Age 70+ years<br>SD N=5<br>SDSD N=508 |
| **Excluded**<br>N=0 baseline serostatus positive/NA<br>N=0 single dose recipients<br>N=0 vaccine error<br>N=0 cases ≤7 days since PB28<br>N=0 follow-up ≤7 days since PB28<br>N=1 samples not collected or out of PB28 window | **Excluded**<br>N=1 baseline serostatus positive/NA<br>N=8 single dose recipients<br>N=0 vaccine error<br>N=0 cases ≤7 days since PB28<br>N=0 follow-up ≤7 days since PB28<br>N=3 samples not collected or out of PB28 window | **Excluded**<br>N=23 baseline serostatus positive/NA<br>N=236 single dose recipients<br>N=2 vaccine error<br>N=13 cases ≤7 days since PB28<br>N=10 follow-up ≤7 days since PB28<br>N=79 samples not collected or out of PB28 window | **Excluded**<br>N=32 baseline serostatus positive/NA<br>N=48 single dose recipients<br>N=0 vaccine error<br>N=2 cases ≤7 days since PB28<br>N=2 follow-up ≤7 days since PB28<br>N=0 samples not collected or out of PB28 window | **Excluded**<br>N=25 baseline serostatus positive/NA<br>N=316 single dose recipients<br>N=4 vaccine error<br>N=28 cases ≤7 days since PB28<br>N=21 follow-up ≤7 days since PB28<br>N=110 samples not collected or out of PB28 window | **Excluded**<br>N=0 baseline serostatus positive/NA<br>N=30 single dose recipients<br>N=0 vaccine error<br>N=0 cases ≤7 days since PB28<br>N=0 follow-up ≤7 days since PB28<br>N=2 samples not collected or out of PB28 window | **Excluded**<br>N=2 baseline serostatus positive/NA<br>N=50 single dose recipients<br>N=0 vaccine error<br>N=0 cases ≤7 days since PB28<br>N=0 follow-up ≤7 days since PB28<br>N=0 samples not collected or out of PB28 window | **Excluded**<br>N=22 baseline serostatus positive/NA<br>N=8 single dose recipients<br>N=2 vaccine error<br>N=8 cases ≤7 days since PB28<br>N=6 follow-up ≤7 days since PB28<br>N=14 samples not collected or out of PB28 window | **Excluded**<br>N=29 baseline serostatus positive/NA<br>N=5 single dose recipients<br>N=1 vaccine error<br>N=12 cases ≤7 days since PB28<br>N=5 follow-up ≤7 days since PB28<br>N=4 samples not collected or out of PB28 window |
| **Eligible**<br>N=59<br><br>**Samples processed**<br>N=1 cases<br>N=27 non cases | **Eligible**<br>N=88<br><br>**Samples processed**<br>N=6 cases<br>N=43 non cases | **Eligible**<br>N=1354<br><br>**Samples processed**<br>N=44 cases<br>N=331 non cases | **Eligible**<br>N=91<br><br>**Samples processed**<br>N=3 cases<br>N=75 non cases | **Eligible**<br>N=1794<br><br>**Samples processed**<br>N=96 cases<br>N=602 non cases | **Eligible**<br>N=28<br><br>**Samples processed**<br>N=0 cases<br>N=26 non cases | **Eligible**<br>N=48<br><br>**Samples processed**<br>N=1 cases<br>N=45 non cases | **Eligible**<br>N=453<br><br>**Samples processed**<br>N=9 cases<br>N=141 non cases | **Eligible**<br>N=457<br><br>**Samples processed**<br>N=11 cases<br>N=117 non cases |

**Figure S2. Immune markers measured at day 28 post-second dose, in primary symptomatic, asymptomatic, non-primary cases, NAAT-positive cases and NAAT-negative non-cases**

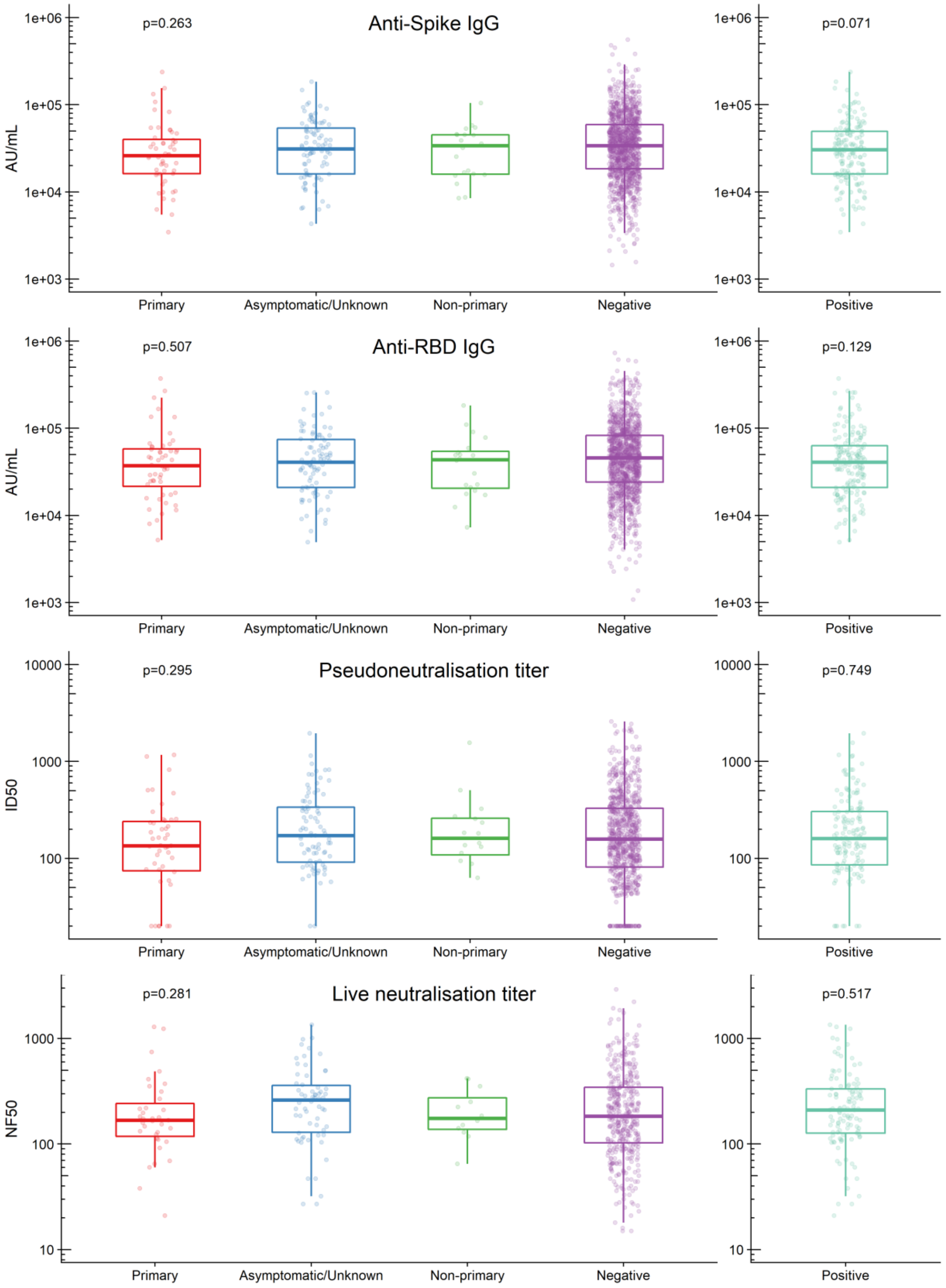


IgG: Immunoglobulin G; RBD: receptor binding domain.

**Figure S3. Correlations between 1). top-left: Anti-SARS-CoV-2 Spike and RBD IgG, 2) top-right: Anti-SARS-CoV-2 Spike IgG and pseudovirus neutralisation titre, 3) bottom-left: Anti-SARS-CoV-2 Spike IgG and live neutralisation titre, 4) bottom-right pseudovirus neutralisation titre and live neutralisation titre.**

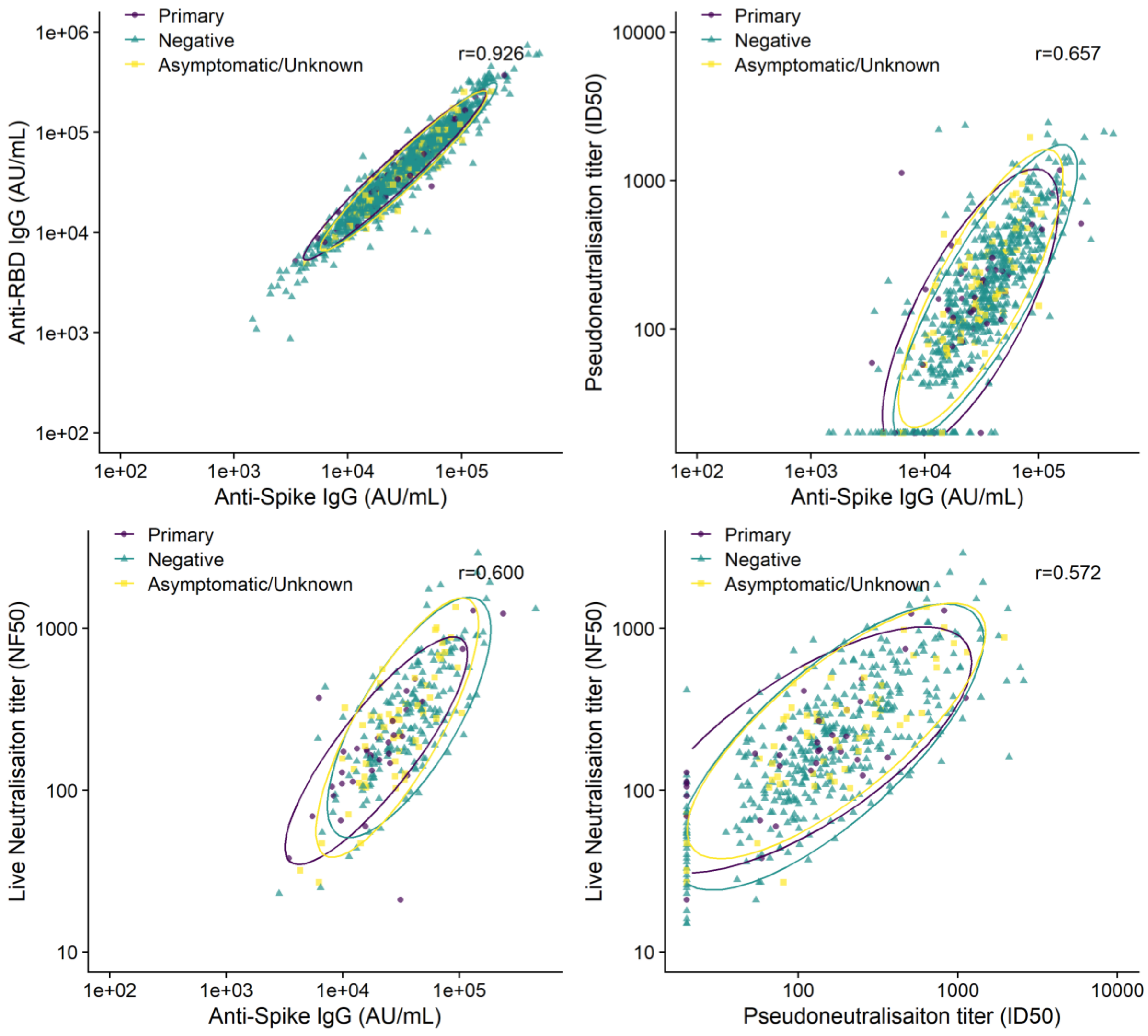


Ellipse shows the 95% confidence intervals for primary symptomatic cases, asymptomatic cases and negative controls assuming t-distribution. Pearson correlation coefficients shown as r values.

**Figure S4A. Adjusted risk of asymptomatic SARS-CoV-2 infection as a function of immune markers measured 28 days post second dose.**

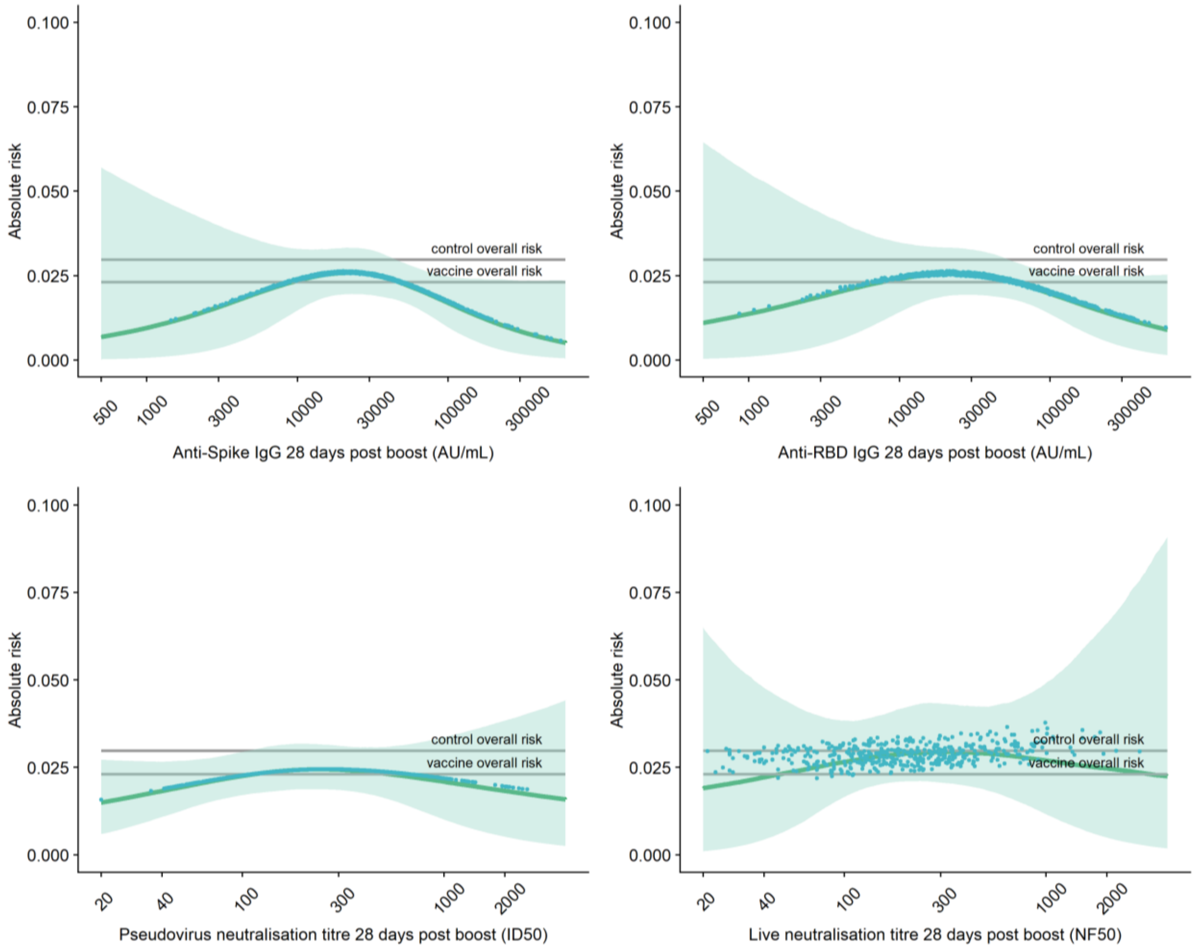


Top left: Anti-Spike IgG 28 days post boost Top right: Anti-RBD IgG 28 days post boost
Bottom left: pseudovirus neutralisation antibody titres 28 days post boost
Bottom right: live neutralisation antibody titres 28 days post boost.
Grey lines show control (MenACWY) overall risk and vaccine (ChAdOx1 nCoV-19) overall risk.
Blue dots show the absolute risk predicted from the model across the range of antibody values included in the analysis, adjusting for baseline exposure risk to SARS-CoV-2 infection (logit-transformed linear covariate including age, ethnicity, BMI, co-morbidities and healthcare worker status). Green shaded areas shows the confidence interval around the predicted mean probability (green line)

**Figure S4B. Sensitivity analysis showing adjusted risk of asymptomatic SARS-CoV-2 infection as a function of immune markers measured 28 days post second dose**

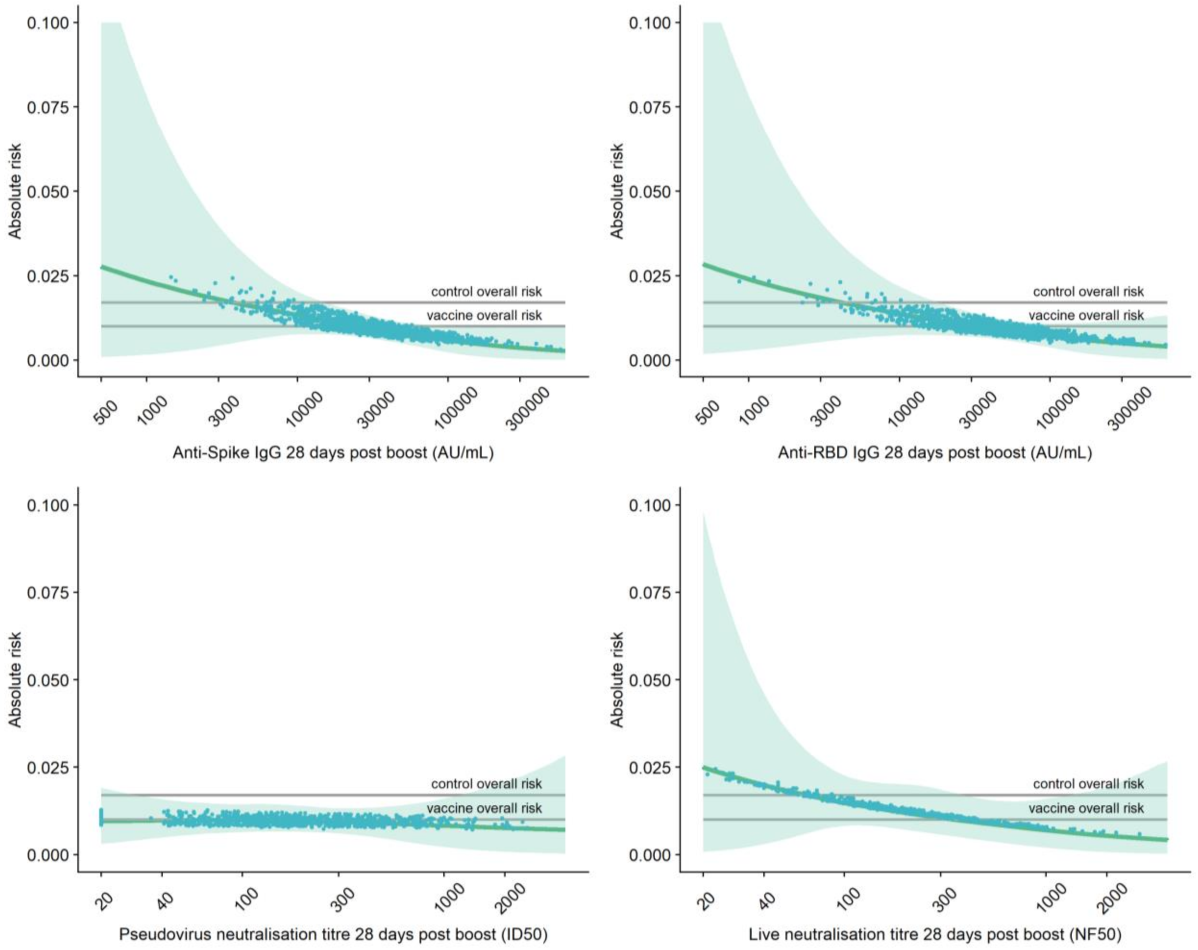


Top left: Anti-Spike IgG 28 days post boost Top right: Anti-RBD IgG 28 days post boost
Bottom left: pseudovirus neutralisation antibody titres 28 days post boost
Bottom right: live neutralisation antibody titres 28 days post boost.
Grey lines show control (MenACWY) overall risk and vaccine (ChAdOx1 nCoV-19) overall risk.
Blue dots show the absolute risk predicted from the model across the range of antibody values included in the analysis, adjusting for baseline exposure risk to SARS-CoV-2 infection (logit-transformed linear covariate including age, ethnicity, BMI, co-morbidities and healthcare worker status). Green shaded areas shows the confidence interval around the predicted mean probability (green line)

**Figure S5A. Adjusted risk of primary symptomatic SARS-CoV-2 infection with shortness of breath as a function of immune markers measured 28 days post second dose**

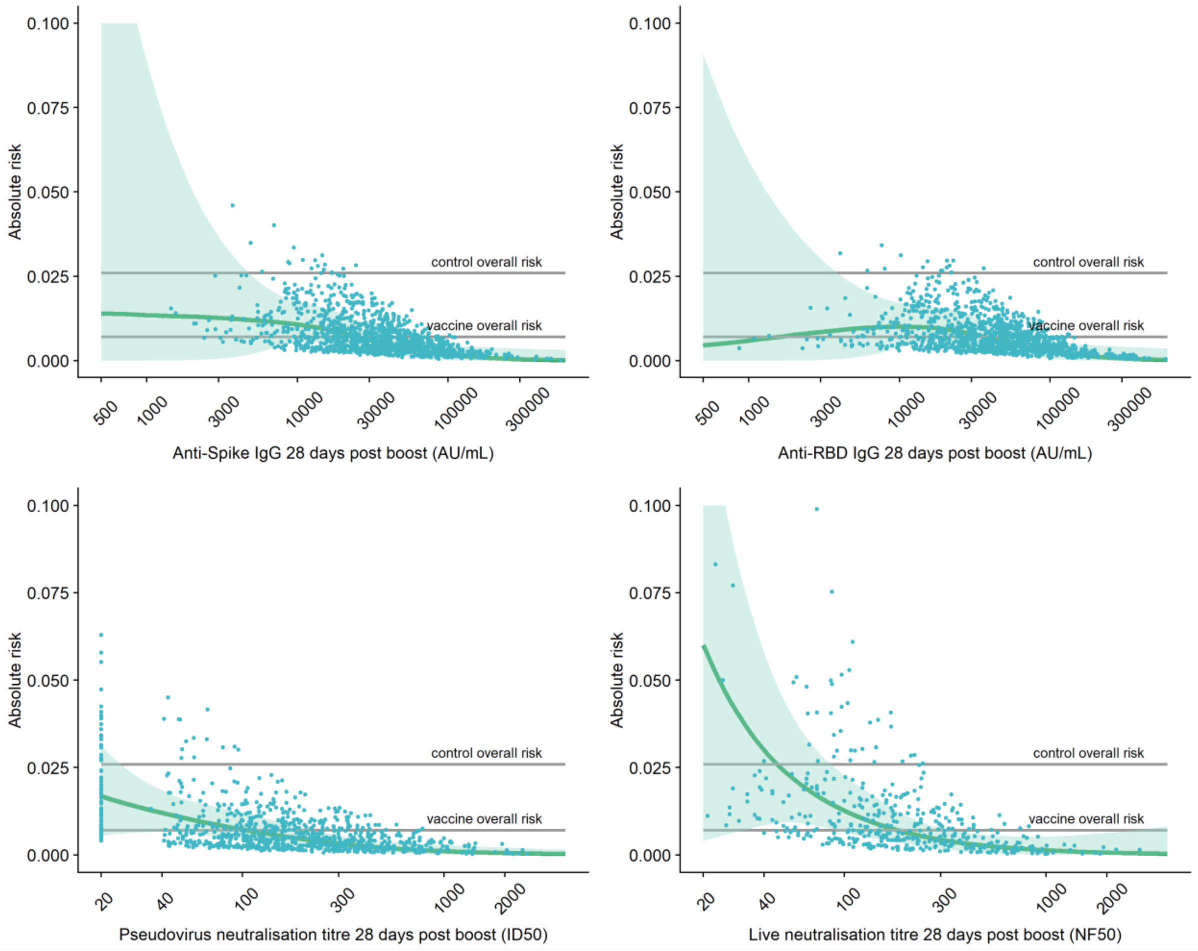


Top left: Anti-Spike IgG 28 days post boost Top right: Anti-RBD IgG 28 days post boost
Bottom left: pseudovirus neutralisation antibody titres 28 days post boost
Bottom right: live neutralisation antibody titres 28 days post boost.
Grey lines show control (MenACWY) overall risk and vaccine (ChAdOx1 nCoV-19) overall risk.
Blue dots show the absolute risk predicted from the model across the range of antibody values included in the analysis, adjusting for baseline exposure risk to SARS-CoV-2 infection (logit-transformed linear covariate including age, ethnicity, BMI, co-morbidities and healthcare worker status). Green shaded areas shows the confidence interval around the predicted mean probability (green line)

**Figure S5B. Adjusted risk of primary symptomatic SARS-CoV-2 infection with no shortness of breath as a function of immune markers measured 28 days post second dose**

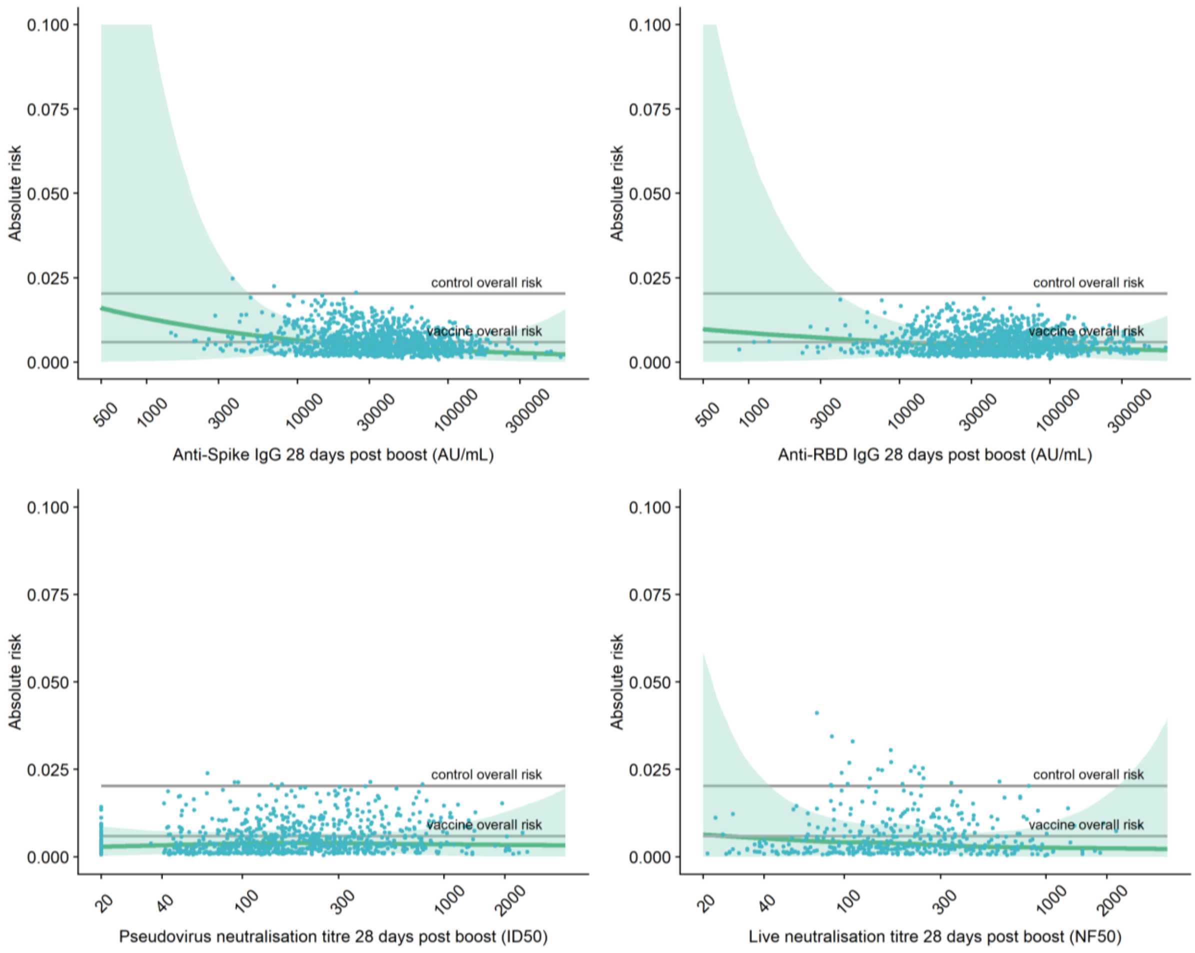


Top left: Anti-Spike IgG 28 days post boost Top right: Anti-RBD IgG 28 days post boost
Bottom left: pseudovirus neutralisation antibody titres 28 days post boost
Bottom right: live neutralisation antibody titres 28 days post boost.
Grey lines show control (MenACWY) overall risk and vaccine (ChAdOx1 nCoV-19) overall risk.
Blue dots show the absolute risk predicted from the model across the range of antibody values included in the analysis, adjusting for baseline exposure risk to SARS-CoV-2 infection (logit-transformed linear covariate including age, ethnicity, BMI, co-morbidities and healthcare worker status). Green shaded areas shows the confidence interval around the predicted mean probability (green line)

**Figure S6A. Adjusted risk of primary symptomatic SARS-CoV-2 infections with 3 or more COVID-19 symptoms as a function of immune markers measured 28 days post second dose**

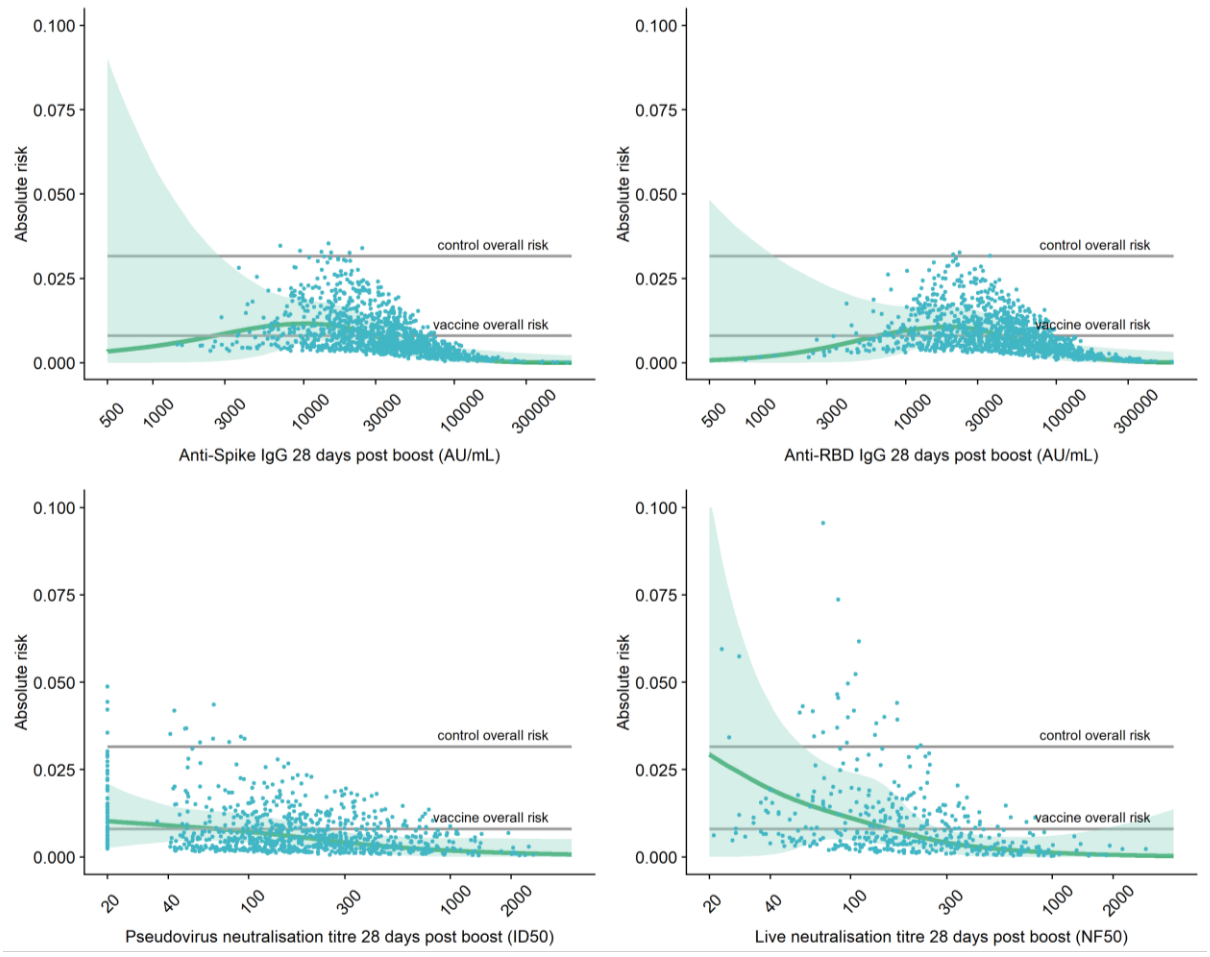


Top left: Anti-Spike IgG 28 days post boost Top right: Anti-RBD IgG 28 days post boost
Bottom left: pseudovirus neutralisation antibody titres 28 days post boost
Bottom right: live neutralisation antibody titres 28 days post boost.
Grey lines show control (MenACWY) overall risk and vaccine (ChAdOx1 nCoV-19) overall risk.
Blue dots show the absolute risk predicted from the model across the range of antibody values included in the analysis, adjusting for baseline exposure risk to SARS-CoV-2 infection (logit-transformed linear covariate including age, ethnicity, BMI, co-morbidities and healthcare worker status). Green shaded areas shows the confidence interval around the predicted mean probability (green line)

**Figure S6B. Relative risk of primary symptomatic SARS-CoV-2 infections with 3 or more COVID-19 symptoms among vaccine recipients compared with MenACWY control arm participants as a function of immune markers measured at day 28 post-second dose**

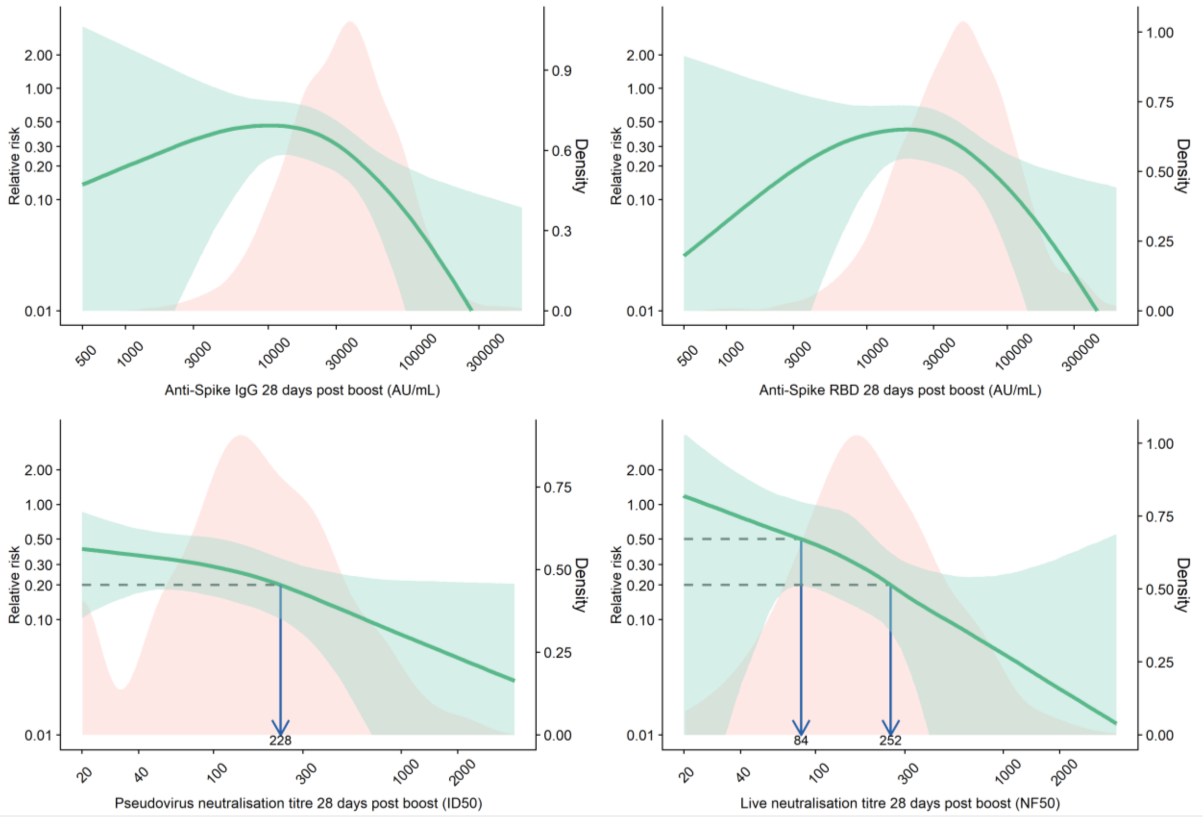


The red shaded areas represent the immune marker density distribution. Green lines show the relative risk of infection among vaccine arm compared to the control arm. Green shaded areas are 95% confidence intervals for the relative risk. The arrows point to the immune marker values at 20% and 50% relative risk, i.e., 80% and 50% vaccine efficacy.

**Table S1. Anti-SARS-CoV-2 Spike and RBD IgG, pseudovirus neutralisation antibody titres and live neutralisation antibody titres (Median and IQR) by primary symptomatic, asymptomatic/unknown, non-primary cases, NAAT positive cases and negative controls**

| Immune marker | Outcome | No. participant | Median (IQR) |
|---|---|---|---|
| **Anti-Spike IgG** | **Negative** | 1155 | 33945 (18450, 59260) |
| | **Positive** | 163 | 30501 (16088, 49529) |
| | Primary | 52 | 26144 (16147, 39996) |
| | Asymptomatic | 91 | 31115 (16112, 54118) |
| | Non-primary | 20 | 33896 (15976, 45307) |
| **Anti-RBD IgG** | **Negative** | 1155 | 45693 (24009, 82432) |
| | **Positive** | 163 | 40884 (20871, 62934) |
| | Primary | 52 | 37276 (21560, 58033) |
| | Asymptomatic | 91 | 40884 (20944, 74226) |
| | Non-primary | 20 | 43673 (20474, 54332) |
| **Pseudovirus neutralisation titre** | **Negative** | 828 | 158 (81, 328) |
| | **Positive** | 149 | 160 (85, 304) |
| | Primary | 47 | 135 (75, 240) |
| | Asymptomatic | 86 | 172 (91, 338) |
| | Non-primary | 16 | 162 (109, 260) |
| **Live neutralisation titre** | **Negative** | 412 | 184 (101, 344) |
| | **Positive** | 110 | 206 (124, 331) |
| | Primary | 36 | 166 (112, 231) |
| | Asymptomatic | 62 | 261 (129, 359) |
| | Non-primary | 12 | 176 (138, 277) |

#### The Oxford Vaccine Trial Group

| | |
|---|---|
| Syed Adlou | Oxford Vaccine Group, Department of Paediatrics, University of Oxford, UK |
| Lauren Allen | National Infection Service, Public Health England, UK |
| Brian Angus | Jenner Institute, Nuffield Department of Medicine, University of Oxford, UK |
| Rachel Anslow | Oxford Vaccine Group, Department of Paediatrics, University of Oxford, UK |
| Marie-Claude Asselin | National Infection Service, Public Health England, UK |
| Natalie Baker | National Infection Service, Public Health England, UK |
| Philip Baker | Oxford Vaccine Group, Department of Paediatrics, University of Oxford, UK |
| Thomas Barlow | National Infection Service, Public Health England, UK |
| Louise Bates | Oxford Vaccine Group, Department of Paediatrics, University of Oxford, UK |
| Amy Beveridge | Oxford Vaccine Group, Department of Paediatrics, University of Oxford, UK |
| Kevin R Bewley | National Infection Service, Public Health England, UK |
| Phillip Brown | National Infection Service, Public Health England, UK |
| Emily Brunt | National Infection Service, Public Health England, UK |
| Karen R Buttigieg | National Infection Service, Public Health England, UK |
| Susana Camara | Oxford Vaccine Group, Department of Paediatrics, University of Oxford, UK |
| Sue Charlton | National Infection Service, Public Health England, UK |
| Emily Chiplin | National Infection Service, Public Health England, UK |
| Paola Cicconi | Jenner Institute, Nuffield Department of Medicine, University of Oxford, UK |
| Elizabeth A Clutterbuck | Oxford Vaccine Group, Department of Paediatrics, University of Oxford, UK |
| Andrea M. Collins | Department of Clinical Sciences, Liverpool School of Tropical Medicine and Liverpool University Hospitals NHS Foundation Trust, Liverpool, UK |
| Naomi S. Coombes | National Infection Service, Public Health England, UK |
| Sue Ann Costa Clemens | Oxford Vaccine Group, Department of Paediatrics, University of Oxford, UK and the Institute of Global Health, University of Siena, Italy |
| Melanie Davison | National Infection Service, Public Health England, UK |
| Tesfaye Demissie | Oxford Vaccine Group, Department of Paediatrics, University of Oxford, UK |
| Tanya Dinesh | Oxford Vaccine Group, Department of Paediatrics, University of Oxford, UK |
| Alexander D. Douglas | Jenner Institute, Nuffield Department of Medicine, University of Oxford, UK |
| Christopher J. A. Duncan | Department of Infection and Tropical Medicine, Newcastle upon Tyne Hospitals NHS Foundation Trust and the Translational and Clinical Research Institute, Immunity and Inflammation Theme, Newcastle University, UK |
| Katherine R. W. Emary | Oxford Vaccine Group, Department of Paediatrics, University of Oxford, UK |
| Katie J. Ewer | Jenner Institute, Nuffield Department of Medicine, University of Oxford, UK |
| Sally Felle | Oxford Vaccine Group, Department of Paediatrics, University of Oxford, UK |
| Daniela M Ferreira | Department of Clinical Sciences, Liverpool School of Tropical Medicine, UK |
| Adam Finn | School of Population Health Sciences, University of Bristol and University Hospitals Bristol and Weston NHS Foundation Trust, UK |
| Pedro M. Folegatti | Jenner Institute, Nuffield Department of Medicine, University of Oxford, UK |
| Ross Fothergill | National Infection Service, Public Health England, UK |
| Sara Fraser | National Infection Service, Public Health England, UK |
| Harriet Garlant | National Infection Service, Public Health England, UK |
| Laura Gatcombe | National Infection Service, Public Health England, UK |
| Kerry J. Godwin | National Infection Service, Public Health England, UK |

| | |
|---|---|
| Anna L Goodman | Department of Infectious Diseases, Guy's and St Thomas' NHS Foundation Trust, St Thomas' Hospital, London, UK and MRC Clinical Trials Unit at University College London, UK |
| Christopher A. Green | NIHR/Wellcome Trust Clinical Research Facility, University Hospitals Birmingham NHS Foundation Trust and Institute of Microbiology & Infection, University of Birmingham, UK |
| Bassam Hallis | National Infection Service, Public Health England, UK |
| Thomas C. Hart | Oxford Vaccine Group, Department of Paediatrics, University of Oxford, UK |
| Paul T. Heath | St George's Vaccine Institute, St George's, University of London, UK |
| Helen Hill | Department of Clinical Sciences, Liverpool School of Tropical Medicine, UK |
| Adrian V. S. Hill | Jenner Institute, Nuffield Department of Medicine, University of Oxford, UK |
| Daniel Jenkin | Jenner Institute, Nuffield Department of Medicine, University of Oxford, UK |
| Mwila Kasanyinga | Oxford Vaccine Group, Department of Paediatrics, University of Oxford, UK |
| Simon Kerridge | Oxford Vaccine Group, Department of Paediatrics, University of Oxford, UK |
| Chanice Knight | National Infection Service, Public Health England, UK |
| Stephanie Leung | National Infection Service, Public Health England, UK |
| Vincenzo Libri | NIHR UCLH Clinical Research Facility and NIHR UCLH Biomedical Research Centre, London, UK |
| Patrick J. Lillie | Hull University Teaching Hospitals NHS Trust and Hull York Medical School, UK |
| Spyridoula Marinou | Oxford Vaccine Group, Department of Paediatrics, University of Oxford, UK |
| Joanna McGlashan | National Infection Service, Public Health England, UK |
| Alastair C. McGregor | London North West University Healthcare NHS Trust and the Department of Medicine, Imperial College London, UK |
| Lorna McInroy | National Infection Service, Public Health England, UK |
| Angela M. Minassian | Jenner Institute, Nuffield Department of Medicine, University of Oxford, UK |
| Yama F Mujadidi | Oxford Vaccine Group, Department of Paediatrics, University of Oxford, UK |
| Elizabeth J. Penn | National Infection Service, Public Health England, UK |
| Katrina M. Pollock | NIHR Imperial Clinical Research Facility and NIHR Imperial Biomedical Research Centre, London, UK |
| Pamela C. Proud | National Infection Service, Public Health England, UK |
| Samuel Provstgaard-Morys | Oxford Vaccine Group, Department of Paediatrics, University of Oxford, UK |
| Durga Rajapaksa | National Infection Service, Public Health England, UK |
| Maheshi N Ramasamy | Oxford Vaccine Group, Department of Paediatrics, University of Oxford, UK |
| Katherine Sanders | Oxford Vaccine Group, Department of Paediatrics, University of Oxford, UK |
| Imam Shaik | National Infection Service, Public Health England, UK |
| Nisha Singh | Oxford Vaccine Group, Department of Paediatrics, University of Oxford, UK |
| Andrew Smith | College of Medical, Veterinary & Life Sciences, Glasgow Dental Hospital & School, University of Glasgow, UK |
| Matthew D. Snape | Oxford Vaccine Group, Department of Paediatrics, University of Oxford, UK |
| Rinn Song | Oxford Vaccine Group, Department of Paediatrics, University of Oxford, UK |
| Sonu Shrestha | Oxford Vaccine Group, Department of Paediatrics, University of Oxford, UK |
| Rebecca K. Sutherland | Clinical Infection Research Group, Regional Infectious Diseases Unit, NHS Lothian, Edinburgh, UK |
| Emma C. Thomson | MRC - University of Glasgow Centre for Virus Research & Department of Infectious Diseases, Queen Elizabeth University Hospital, Glasgow, UK |
| David P. J. Turner | University of Nottingham and Nottingham University Hospitals NHS Trust, UK |
| Alice Webb-Bridges | Oxford Vaccine Group, Department of Paediatrics, University of Oxford, UK |
| Christopher J Williams | Aneurin Bevan University Health Board, Newport, Wales |

#### Acknowledgements

| Clinical Trials Research Governance Office, University of Oxford |
|---|
| Ronja Bahadori |
| Elaine Chick |
| Heather House |
| Claire Riddle |
| **Data and Safety Monitoring Board (DSMB)** |
| George Bouliotis |
| Steve Black |
| Elizabeth Bukusi |
| Cornelia Dekker |
| Robert Heyderman |
| Gregory Hussey |
| Paul Kaye |
| Bernhards Ogutu |
| Walter Orenstein |
| Sonia Ramos |
| Manish Sadarangani |
| **Department of Paediatrics, University of Oxford** |
| Georg A. Holländer |
| **Endpoint Evaluation Committee** |
| Jeremy Carr |
| Steve Chambers |
| Kim Davis |
| Simon Drysdale |
| Malick Gibani |
| Elizabeth Hammershaimb |
| Michael Harrington |

| |
|---|
| Celina Jin |
| Seilesh Kadambari |
| Rama Kandasamy |
| Toby Maher |
| Jamilah Meghji |
| Claire Munro |
| David Pace |
| Rekha Rapaka |
| Robindra Basu Roy |
| Daniel Silman |
| Gemma Sinclair |
| Jing Wang |
| **Jenner Institute, University of Oxford** |
| Iona Tarbet |
| **Nuffield Department of Medicine, University of Oxford** |
| Richard Cornall |
| Richard Liwicki |
| Denis Murphy |
| Elizabeth Salter |
| Katherine Skinner |
| Philip Taylor |
| Oto Velicka |
| **Oxford Research Services (Contracts)** |
| Carly Banner |
| Sally Pelling-Deeves |
| Gary Priest |
| **Oxford University Hospitals Trust** |
| Bruno Holthof |
| **The Oxford COVID-19 Vaccine Trial Team** |
| **Public Affairs Directorate and Divisional Communication Team** |
| Alison Brindle |

| |
|---|
| Alexander Buxton |
| James Colman |
| Chris McIntyre |
| Steve Pritchard |
